# Lifestyle therapy versus cognitive behavioural therapy for adults with mood disorders: a randomised non-inferiority trial

**DOI:** 10.64898/2026.09.08.26362466

**Authors:** Adrienne O’Neil, Karyn Richardson, Jessica A Davis, Tayla John, Sophie Mahoney, Marita Bryan, Rachel Fiddes, Denny Meyer, Dean Saunders, Mary Lou Chatterton, Megan Turner, Sally Brown, Felice N Jacka, Niamh L Mundell, Marlene Gojanovic, Lara K Radovic, Eileen McDonald, Madeleine L. Connolly, Greg Murray, Neil Thomas, Sanna Barrand, Melissa O’Shea, Tetyana Rocks, Elena S. George, Wolfgang Marx, Md Parvez Mosharaf, Tabinda Jabeen, Anu Ruusunen, Ravi Iyer, Cathrine Mihalopoulos, Michael Berk

## Abstract

**Objective:** To determine whether a multicomponent lifestyle intervention delivered by accredited dietitians and exercise physiologists was non-inferior to cognitive behavioural therapy (CBT) for reducing depressive symptoms in adults with moderate-to-severe mood disorders.

**Design:** Randomised, parallel-group, single-blind, non-inferiority trial.

**Setting:** Community-based recruitment across Australia with interventions delivered via telehealth.

**Participants:** 358 adults with major depressive disorder or bipolar depression and moderate-to-severe depressive symptoms.

**Interventions:** Lifestyle therapy versus brief structured CBT.

**Main outcome measures:** Blinded interviewer-rated MADRS at 8 weeks with a prespecified non-inferiority margin of −1.6 points.

**Results:** Mean session attendance was 5.11/7 in the lifestyle therapy group and 5.40/7 in the CBT group; 91% of participants attended the initial individual session, and primary outcome data were available for 267 participants (130 lifestyle therapy; 137 CBT). Non-inferiority was supported at 8 weeks in both the intention-to-treat (β=1.76, 95% CI −0.28 to 3.81) and per-protocol (β=1.58, 95% CI −0.56 to 3.73) analyses. Depressive symptoms decreased in both groups. Mean MADRS scores decreased by 8.55 points (95% CI 7.09 to 10.02) with lifestyle therapy and 6.82 points (5.38 to 8.25) with CBT in the intention-to-treat analysis, with similar findings in the per-protocol analysis (8.37 points [6.84 to 9.90] and 6.79 points [5.27 to 8.31], respectively).

**Conclusions:** Lifestyle therapy was non-inferior to CBT and may expand access to evidence-based depression care through a broader allied health workforce.

**Trial Registration:** Australian New Zealand Clinical Trials Registry (Registry number: ACTRN12622001026718) https://www.anzctr.org.au/

**What is already known on this topic:**

- Lifestyle interventions are recommended in international clinical guidelines for adults with mood disorders.
- Most previous randomised trials have compared lifestyle interventions with usual care or inactive controls rather than established psychological treatments.
- Whether lifestyle therapy achieves clinical outcomes comparable to cognitive behavioural therapy (CBT) is uncertain.

**What this study adds:**

- In this randomised non-inferiority trial, lifestyle therapy delivered by accredited dietitians and exercise physiologists was non-inferior to a brief structured CBT programme for reducing depressive symptoms in adults with moderate-to-severe mood disorders.
- Lifestyle therapy also improved several health behaviours, including diet quality, sleep, and alcohol and substance use risk.
- Appropriately trained dietitians and exercise physiologists can deliver a first-line depression treatment producing outcomes comparable with psychologist-delivered CBT.

## INTRODUCTION

Mood disorders are among the leading causes of disability worldwide^1^, and demand for mental health care continues to outpace the capacity of existing services. Mental health workforce shortages have become a major barrier to delivering timely, guideline-concordant care, highlighting the need for effective interventions that can be delivered by a broader clinical workforce.

Lifestyle therapies are increasingly recommended in clinical guidelines for mood disorders in United Kingdom^2^, Canada^3^, Europe^4^, United States^5^ and Australasia^6^; however, implementation in routine mental health care remains limited^7^. These recommendations reflect growing evidence that interventions targeting diet, physical activity, sleep, and substance use can improve depressive symptoms. One important translational barrier is the absence of evidence demonstrating whether lifestyle therapy achieves outcomes comparable to established first-line psychological treatments. Identifying interventions that can be safely and effectively delivered by existing allied health workforces is therefore an international health system priority.

To date, no randomised non-inferiority trial has evaluated whether a multicomponent lifestyle intervention is non-inferior to cognitive behavioural therapy (CBT) in adults with moderate-to-severe, clinically diagnosed mood disorders. Such evidence is needed to inform clinical decision-making, workforce planning, and implementation of guideline recommendations, particularly where lifestyle interventions are already endorsed as a foundational component of care^6^. Demonstrating comparable effectiveness would also support more personalised treatment pathways by providing an evidence-based alternative for people who prefer lifestyle-focused care or who experience barriers to accessing psychological therapy.

To address this gap, we conducted the HARMON-E randomised non-inferiority trial comparing a multicomponent lifestyle intervention delivered by accredited dietitians and exercise physiologists with a brief, structured group CBT programme delivered by psychologists. Both interventions were delivered via telehealth using matched treatment intensity and group format. We hypothesised that lifestyle therapy would be non-inferior to CBT in reducing interviewer-rated depressive symptoms after eight weeks of treatment in adults with moderate-to-severe unipolar or bipolar depression.

## METHODS

### Study design

The HARMON-E trial was an investigator-initiated, two-arm, parallel, individually randomised, group treatment, non-inferiority trial conducted online via videoconference across Australia. Involvement of lived experience research partners in the research team and steering committee informed the trial design, methods and processes. Both arms were matched for frequency, intensity and scheduling (i.e., seven sessions over 8 weeks [1 one-on-one, 6 group sessions). The trial protocol was published previously^8^ and approved by Deakin University (2022-273) and Barwon Health Human Research Ethics Committees (HREC)(22/48). The trial was conducted in accordance with the Declaration of Helsinki, Australian National Health and Medical Research Council National Statement on Ethical Conduct in Human Research (2007) and ICH Guidelines for Good Clinical Practice. All participants provided written informed consent. This manuscript adheres to CONSORT Statements for reporting non-inferiority and equivalence trials and non-pharmacological interventions.

### Participants

Participants were recruited via community and Meta advertisements. Eligible participants were 18+ years, Australia-based, meeting the following criteria: moderate-to-severe depression (Montgomery-Åsberg Depression Rating Scale (MADRS) ≥ 20) experiencing/had experienced a major depressive episode in the past 2-years, confirmed using the Structured Clinical Interview for DSM-5 Research Version (SCID-5 RV); could provide informed consent and converse in English; were clinically suited to online group-based therapy; had access to a computer and internet literacy. Participants were excluded if they had optimal lifestyle behaviours; severe food allergies, intolerances, aversions or malabsorption issues; were medically unfit to engage in general group exercise; pregnant, breastfeeding, or planning pregnancy within 12-months; had socio-cultural, religious, or medical contraindications to lifestyle interventions; a known/suspected clinically unstable systemic medical disorder; participation in another intervention study; a current, or past diagnosis and/or treatment for an eating disorder; commenced a new/duplicate treatment for mental illness in the last 30 days; experienced an episode of hypomania/mania in the last 30 days; or received treatment for an acute mental health concern (inpatient setting/emergency department) within the last 30 days.

### Randomisation and masking

Randomisation occurred after baseline assessment, once four participants had been enrolled in each arm. Participants were randomised to lifestyle or psychotherapy (1:1) using computer-generated randomisation in blocks of 8 by an independent statistician. The unblinded Clinical Trial Coordinator informed participants of their allocation. This was a single-blind trial; investigators, data assessors and the study statistician were blind.

Participants and interventionists could not be blinded. Participants were instructed to conceal their allocation from blinded assessors.

### Procedures

Advertisements directed potential participants to a pre-screening survey in REDCap, assessing likely eligibility based on: (1) age, (2) location, (3) lifestyle behaviours, (4) disordered eating and (5) depressive symptoms (using a stepped screening approach). Eligible participants underwent a clinical interview via videoconference to confirm mood disorder status: (1) current or major depressive episode in the past 2-years (SCID-5 RV) and (2) moderate-to-severe depressive symptoms (MADRS≥ 20 for unipolar or bipolar depression)^9^. Baseline and primary endpoint assessments were conducted by blinded research staff via videoconference and 16- and 26-week follow-ups (PHQ-9) were self-reported via email and SMS. Both programs were manualised. All participants received intervention-specific workbooks and hampers. Sessions were recorded; 10% were independently assessed for treatment fidelity.

### Experimental condition (lifestyle therapy)

Both interventions were intentionally matched for treatment dose, delivery format, session frequency, duration, and opportunities for participant interaction. Each intervention comprised an individual introductory session followed by six therapist-led group sessions delivered via telehealth over eight weeks. This design sought to ensure that the principal difference between study arms was the therapeutic content rather than treatment intensity, clinician contact, or group support, thereby providing a rigorous comparison of two active treatment approaches.

The lifestyle intervention has been described elsewhere^8^. It was adapted from the CALM and SMILES trials^10^ ^11^, which targeted nutrition and physical activity, expanded here to include sleep quality and substance use, aligned with clinical guidelines^6^. The program was co-delivered with accredited dietitians and exercise physiologists supported by a mental health nursing consultation and escalation framework. Participants were provided with relevant information on lifestyle for mood disorders, goal-based activities such as creating balanced meals (using a weight-neutral approach), key nutrients and foods for mental health, mindful eating, finding enjoyable physical activity across different contexts, practical examples of physical activity, and tips for sleep and alcohol management (Supplementary Table 1).

### Comparator control condition (psychotherapy)

The psychotherapy intervention comprised a brief, manualised cognitive behavioural therapy (CBT) programme adapted from the validated *Mood Management Course*, an evidence-based group CBT intervention for depression. The programme was designed to reduce depressive symptoms by helping participants identify and modify maladaptive patterns of thinking and behaviour through structured CBT skill development. Sessions were co-facilitated by either two registered psychologists or a senior clinical psychologist and a psychologist-in-training. Core CBT components included psychoeducation, behavioural activation, identification of cognitive distortions, cognitive restructuring, problem-solving, relapse prevention, and structured between-session practice. Participants were encouraged to apply CBT strategies between sessions through homework exercises, with progress reviewed during subsequent sessions. Session content followed a standardised treatment manual (Supplementary Table 1). To preserve treatment fidelity and minimise contamination between study arms, facilitators did not provide counselling or advice regarding diet, physical activity, sleep, or substance use, which constituted the active components of the lifestyle intervention.

### Outcomes

Primary and secondary outcomes were collected at baseline and 8-week follow-up, and additional PHQ-9 outcomes at 16- and 26 weeks. The primary outcome was the MADRS, a blinded 10-item interviewer-rated assessment of depressive severity over the previous week. Secondary outcomes included mental health, quality of life, functional recovery, and lifestyle related outcomes (see^8^). Achievement of group program goals were assessed within each arm. Group program goals in the lifestyle therapy arm were defined as (i) at least a 15% improvement on “HARMON-E Diet” score (adherence to Mediterranean diet), (ii) reductions in participants’ risk profile category on the Alcohol, Smoking and Substance Involvement Screening Test (ASSIST) questionnaire, (iii) reductions in participants’ risk profile category for at least one substance on the ASSIST questionnaire by 8 weeks, (iv) reductions in participants’ insomnia severity profile category on the Insomnia Severity Index (ISI) questionnaire, and (v) Australian physical activity guidelines of 150 min of moderate intensity or 75 min of vigorous intensity physical activity per week (SIMPAQ and Borg scales) at 8 weeks. Program goals in the psychotherapy arm were assessed using the CBT Suitability Scale (CBT-SUITS) (i) a 10% improvement for total score, (ii) a 10% improvement for the rationale subscale, (iii) a 10% improvement for the insight subscale, and (iv) a 10% improvement for the behaviour subscale. While designed to evaluate attitudes towards and suitability for CBT, CBT-SUITS predicts treatment outcomes; the insight subscale is associated with post-treatment symptom change^12^.

Weekly online questionnaires were used to monitor general distress (Depression Anxiety and Stress Scale (DASS-10)) with safety events also collected at weeks 16 and 26. The seriousness, severity, expectedness, and relatedness of safety events to either intervention were assessed by trial medical personnel and reported to the DSMB, HREC, and sponsor, as appropriate.

#### Sample Size & Statistical Analysis

For the primary hypothesis, a non-inferiority margin was set at -1.6 MADRS points at 8 weeks, in line with the accepted cut-off for detecting a minimal clinically important difference in individuals with mood disorders^9^. The power analysis assumed a pooled standard deviation of 3.6 (baseline variance=4.03) and 80% power, resulting in a total sample size of 126. An Intraclass Correlation Coefficient (ICC) of 0.2 was used to account for variation between arms (average group size=8) and a conservative 20% attrition rate, resulting in a planned sample size of 378 participants. For the secondary outcome of PHQ-9 depression outcomes at 8, 16 and 26 weeks, we set a non-inferiority margin of -2.0 PHQ-9 points^10^.

Because intention-to-treat (ITT) analyses in non-inferiority trials may favour conclusions of treatment similarity, and per-protocol (PP) analyses can reduce type I error, both analytical approaches were undertaken. Confidence in a non-inferiority conclusion is greatest when ITT and PP findings are concordant^13^. ITT analyses included all randomised participants and PP included those with ≥50% session attendance. Baseline imbalances were assessed using chi-square and Mann–Whitney tests, and variables associated with attrition using logistic regression. The primary outcome was analysed using linear mixed models comparing change in MADRS scores from baseline to week 8, adjusting for covariates. Effects were reported as coefficients with 95% confidence intervals (CIs). Non-inferiority was concluded when the entire 95% CI for the between-arm difference in change scores remained above the predefined non-inferiority margin. We also presented the percentage of participants meeting criteria for remission (MADRS <10)^14^ at 8 weeks.

Exploratory aims concerned possible arm-moderation effects on changes in MADRS scores, addressed using ANCOVA analyses to test for interaction effects between arm and potential moderators (readiness to change, self-efficacy, personality disorder rating scale, and medication adherence). Sub-group analysis was conducted to test non-inferiority hypothesis separately for those with major depressive disorder (MDD) and bipolar disorder (BD), and to explore longer-term treatment effects on self-reported depressive symptoms using the PHQ-9 at 8, 16 and 26 weeks.

LMM analyses were conducted for the secondary outcomes using the ITT and PP samples (ITT only presented). Due to the large number of variables, Bonferroni adjustment was necessary, with p-values <0.001 considered significant. Finally, 95% CIs were presented separately for each arm when significant improvements were observed.

## RESULTS

Between September 2022 and October 2025, 358 participants were enrolled. Final follow-up was completed in April 2026. Participant flow is shown in Figure 1.

**Figure 1.**
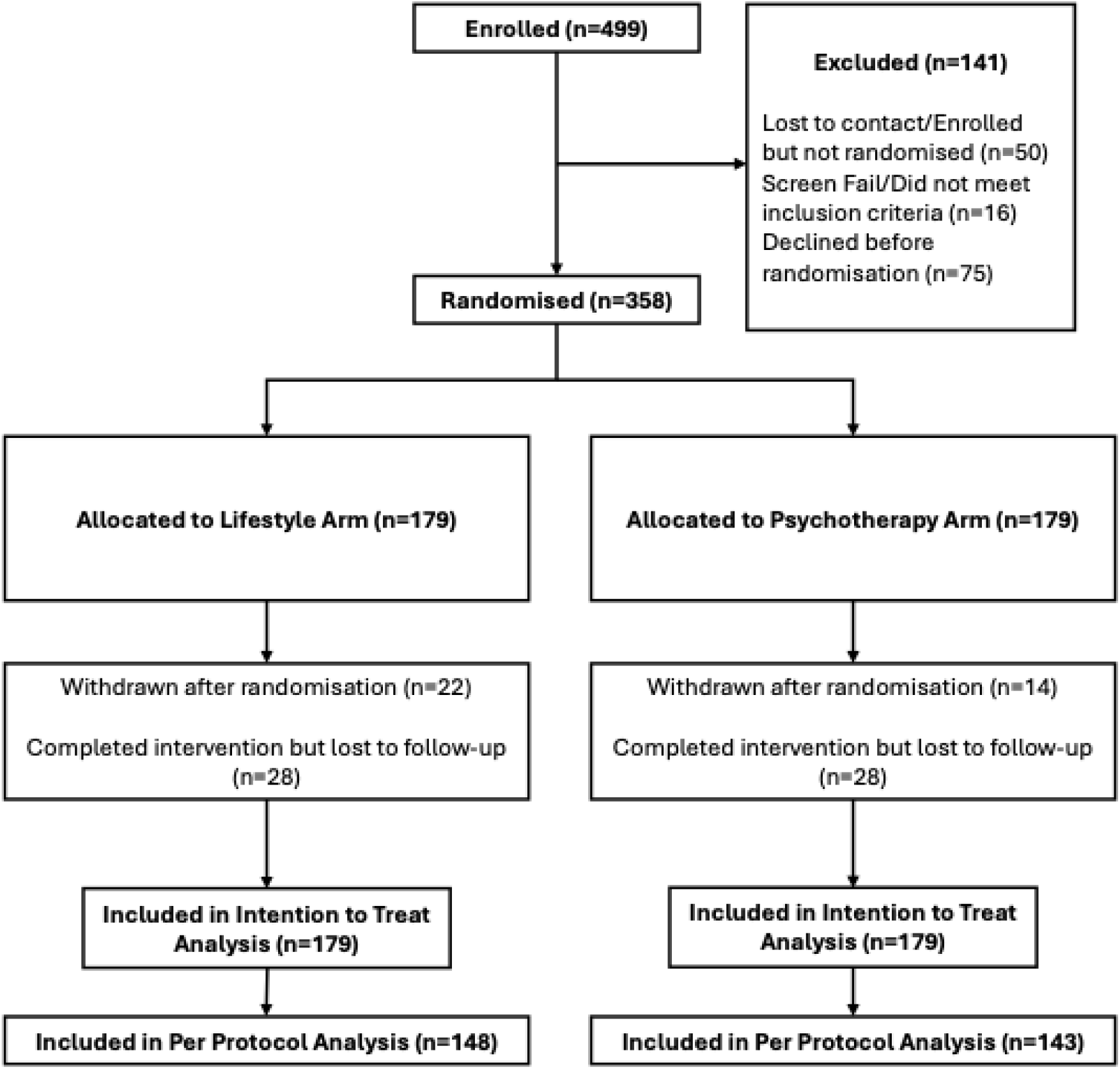
HARMON-E Trial Flowchart

Baseline characteristics are presented in Table 1. Participants had a mean age of 44-years (SD=13.0). Most were women (80%). Mean baseline MADRS score was 26.9 (SD=6.54), indicating moderate depression severity. Nearly 90% were taking psychiatric medication (88.9%).

**Table 1.**
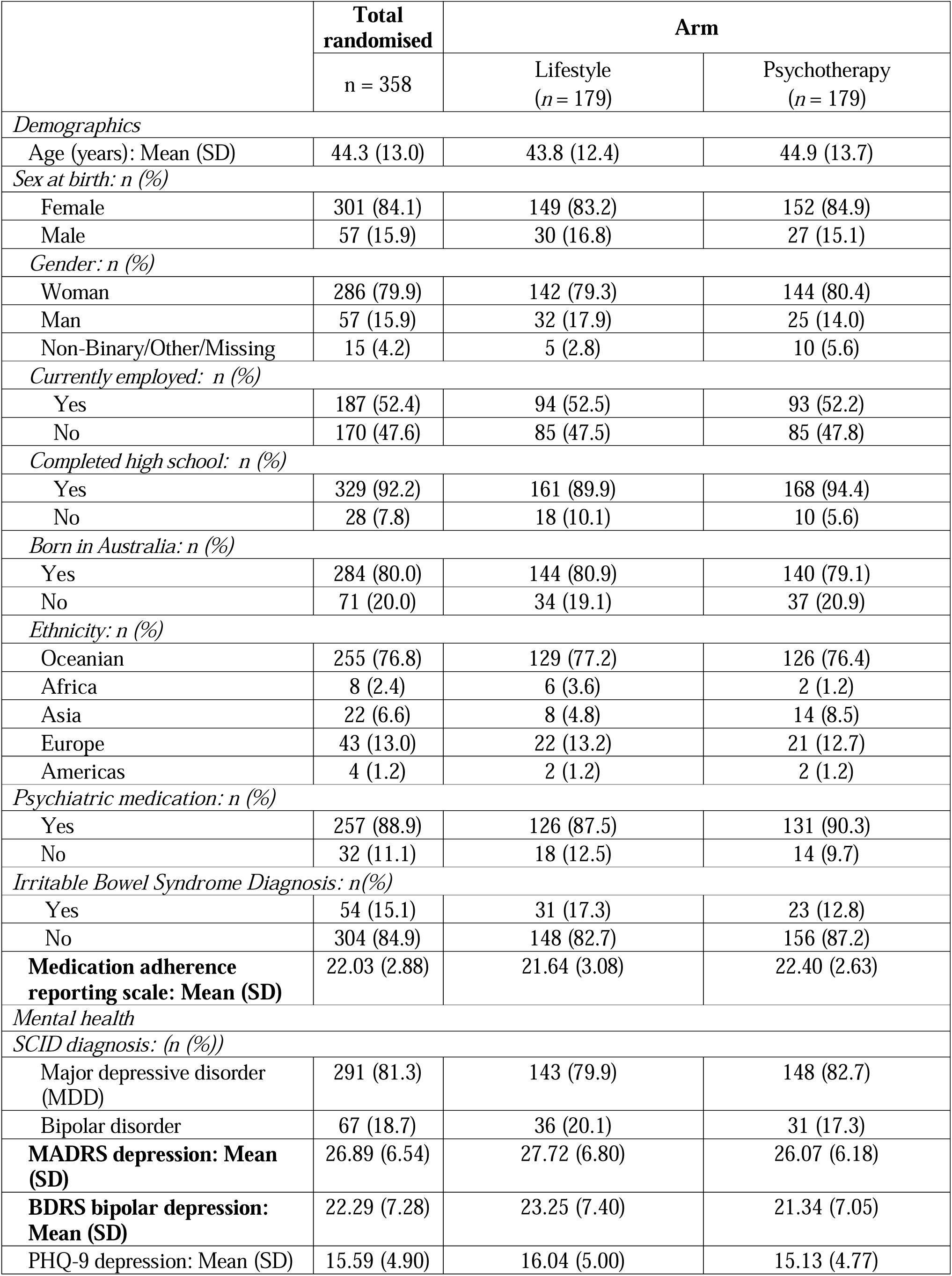
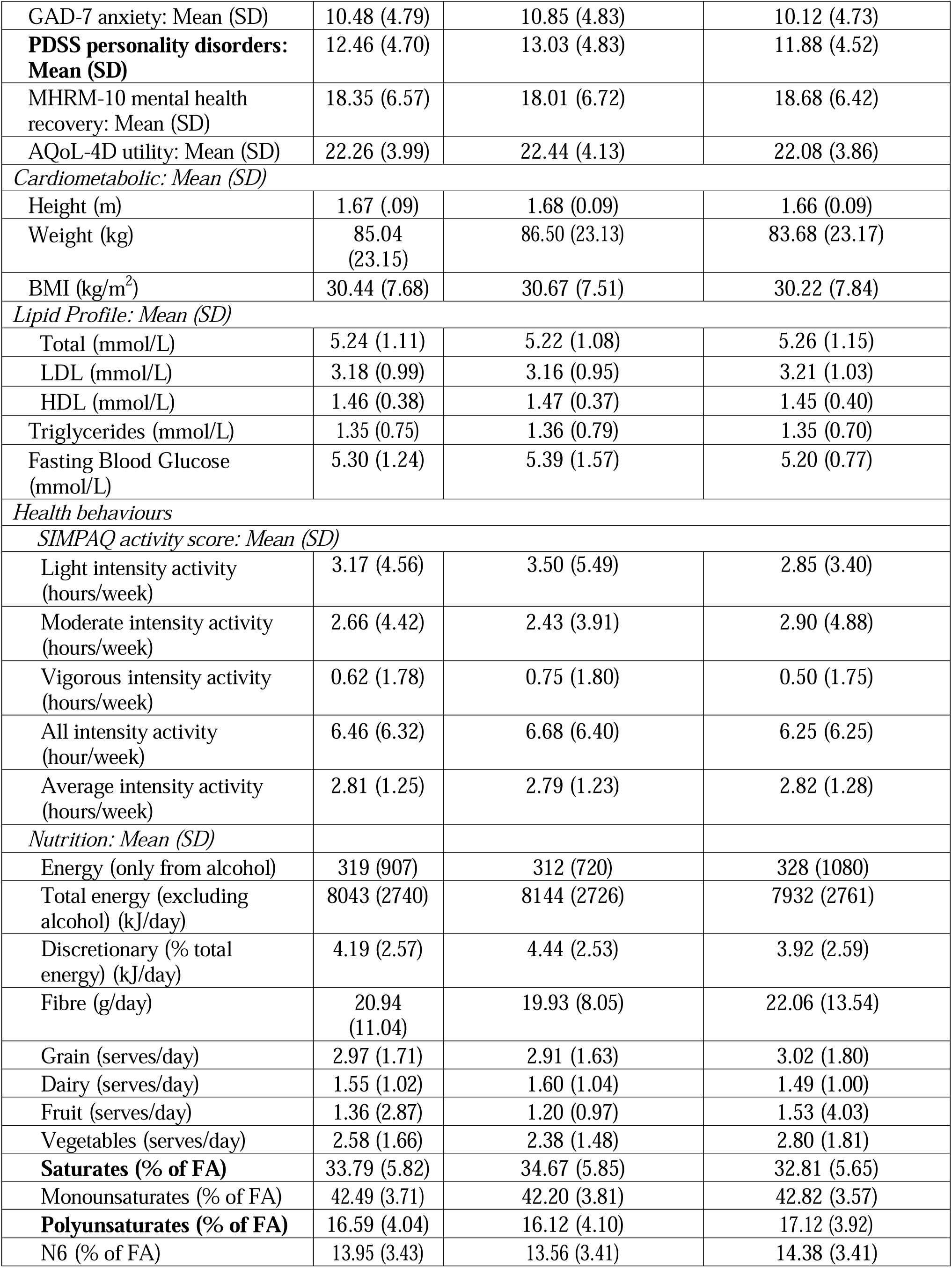
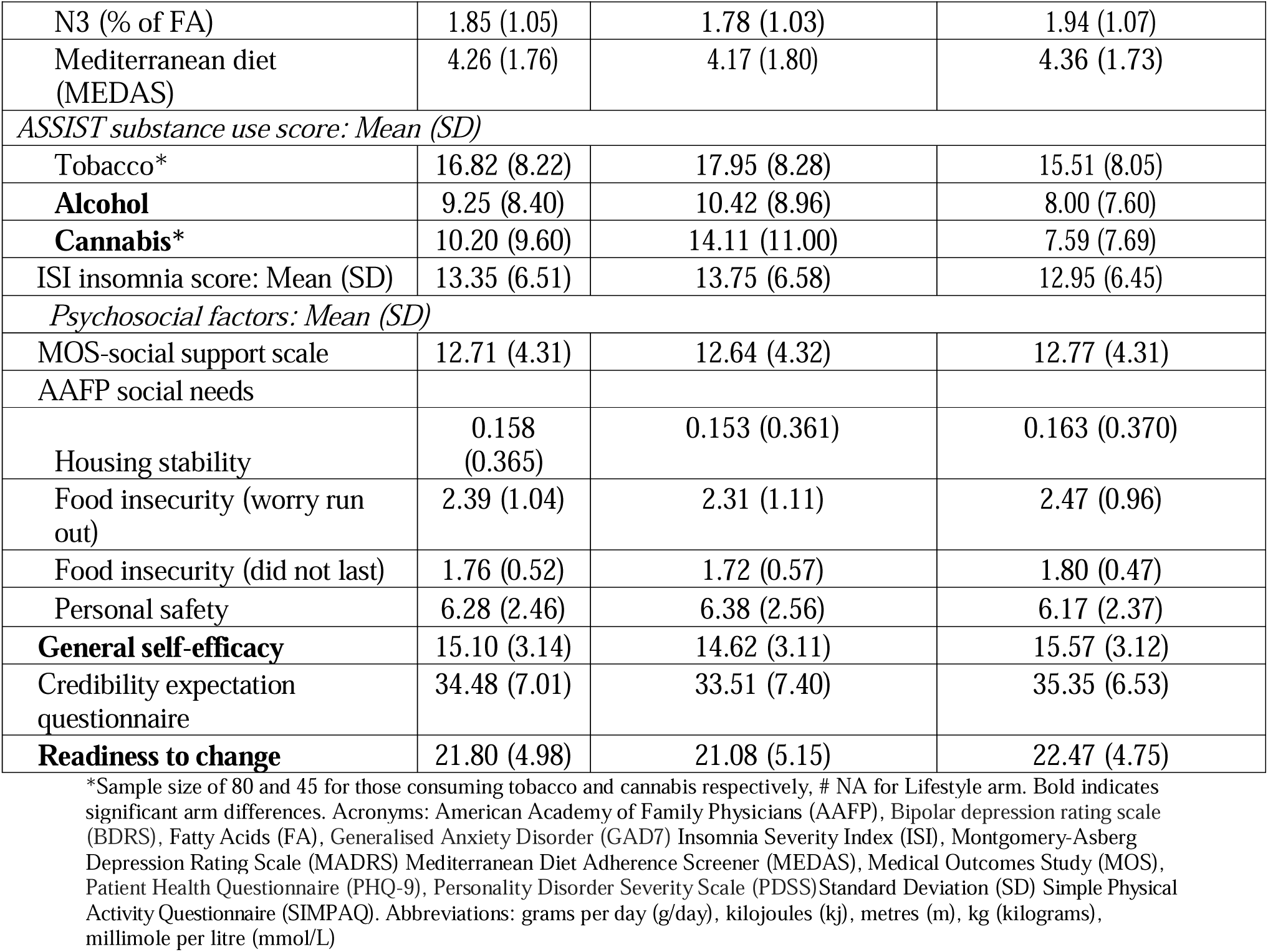
Baseline characteristics of study sample.

### Intervention delivery

Of those commencing treatment, 291 (81.2%) attended 4+ sessions. Mean session attendance was comparable between lifestyle therapy (mean=5.11, SD=2.07) and psychotherapy (mean=5.40, SD=2.06). Fidelity assessments demonstrated high adherence to intervention protocols (Table S2–3)(Lifestyle =93.7%; Psychotherapy=92.0%).

At the primary endpoint (week 8), attrition in the ITT sample was 25.4%, with no significant difference between lifestyle therapy and psychotherapy (27.4% vs 23.5%, p=0.395). Mean age of dropouts 40.6 years compared to 45.6 years among completers. No baseline clinical characteristics predicted attrition. Attrition was lower within the per-protocol sample (12.4%) and similar between arms (lifestyle: 13.3% vs psychotherapy: 11.5%).

### Primary outcome

The 95% CI for the between-group difference in change scores (β= 1.76; 95% CI -0.28-3.81 MADRS points) remained entirely above the prespecified non-inferiority margin of −1.6 points, supporting non-inferiority (Table 3). No significant treatment-by-time interaction was observed (F(1,320)=2.89, p=.090), indicating comparable symptom trajectories between arms (Figure 2). Results were consistent in the PP analysis (β=1.58; 95% CI: -0.56-3.73, n=291). Depressive symptoms improved in both arms. Over 8 weeks, MADRS scores decreased by 31% within the ITT sample in lifestyle therapy participants and 26% in psychotherapy participants with mean reductions of 8.55 points (95% CI: 7.09-10.02) and 6.82 (95% CIs: 5.38-8.25), respectively (Table 2). Within the per-protocol sample, mean reductions were 8.37 points (95% CI: 6.84–9.90) and 6.79 points (95% CI: 5.27–8.31), respectively. The percentage of participants who achieved remission was 21.7% and 17.6% in the lifestyle and psychotherapy arm, respectively (ITT sample) (p=0.406).

**Figure 2.**
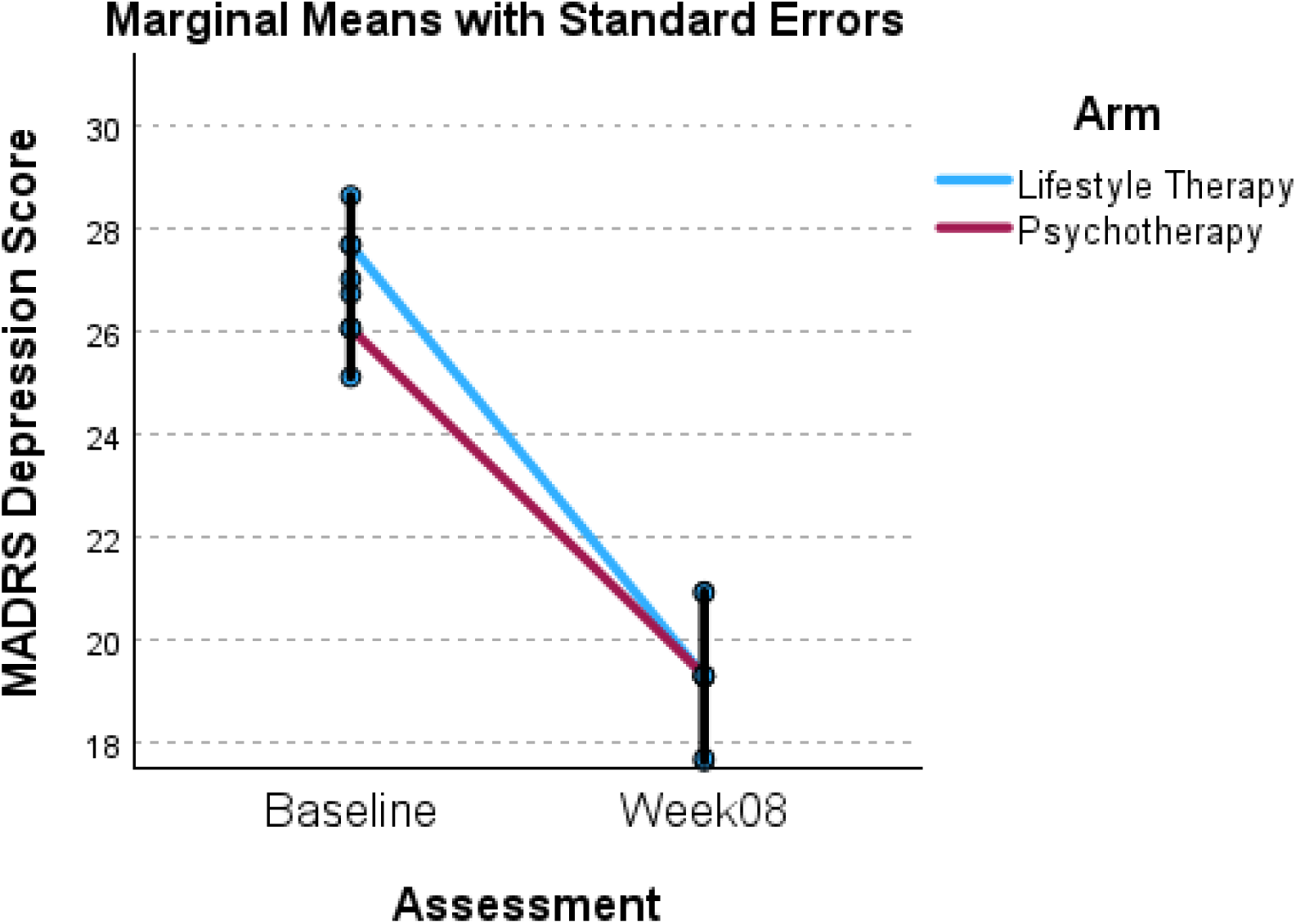
Intention To Treat analysis for MADRS over 8 weeks controlling for age at enrolment

**Table 2.**
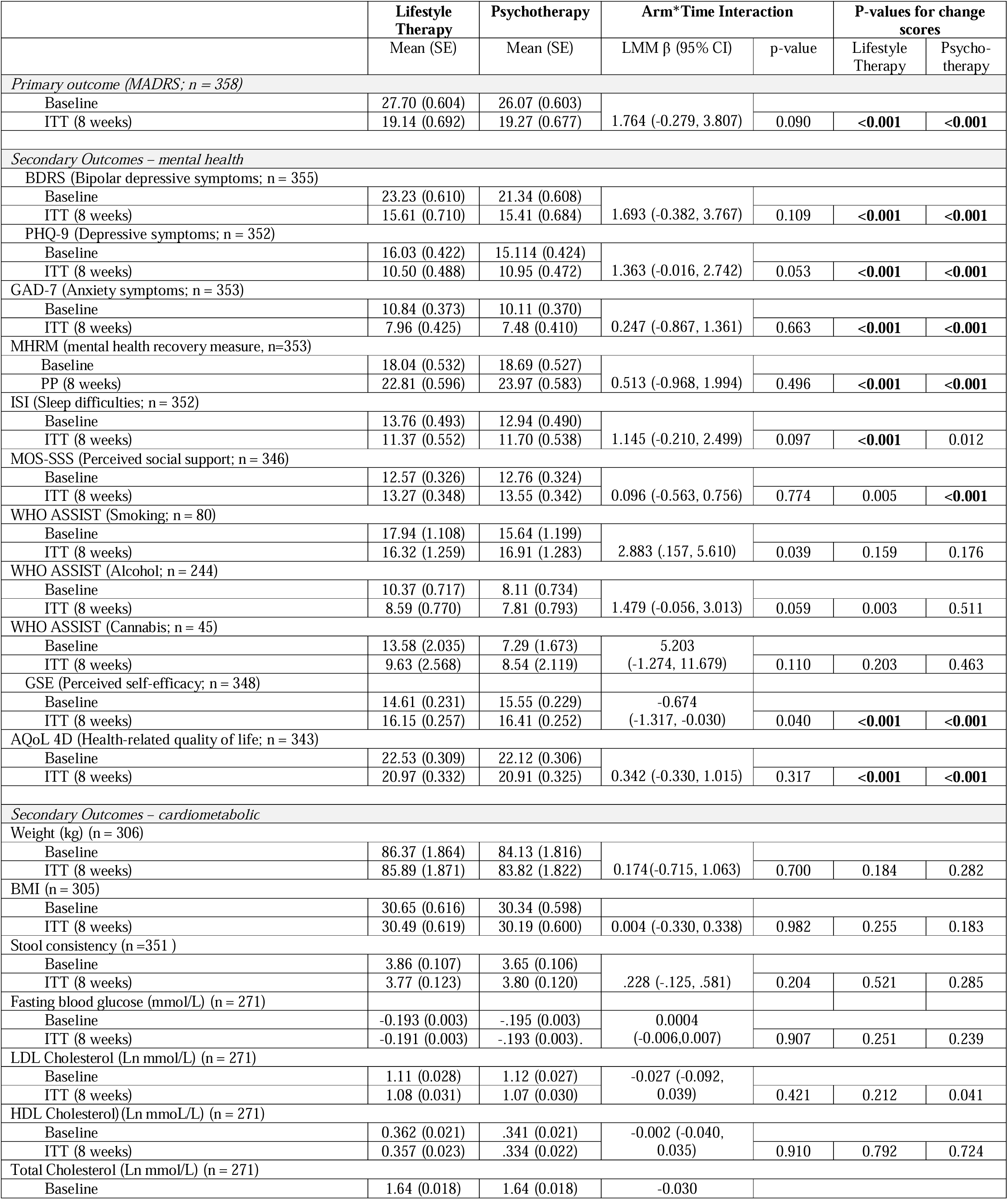
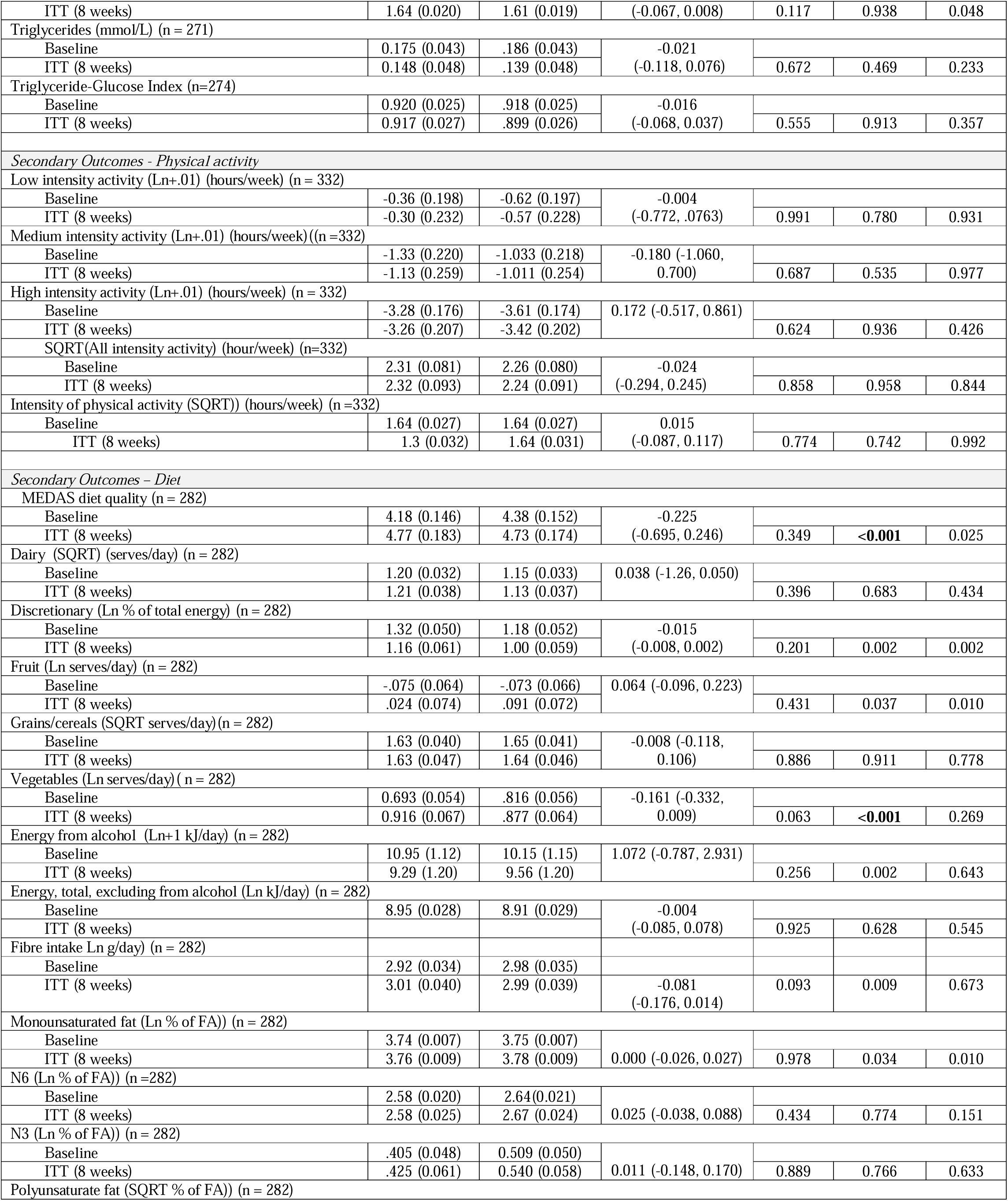
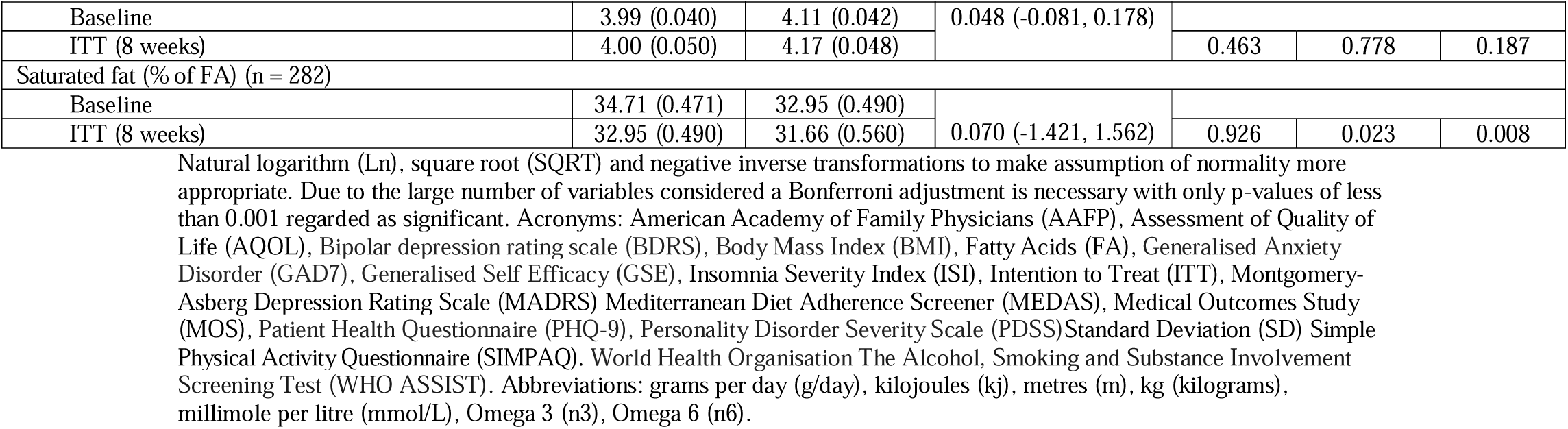
Primary and Secondary Outcomes using LMM.

**Table 3.**
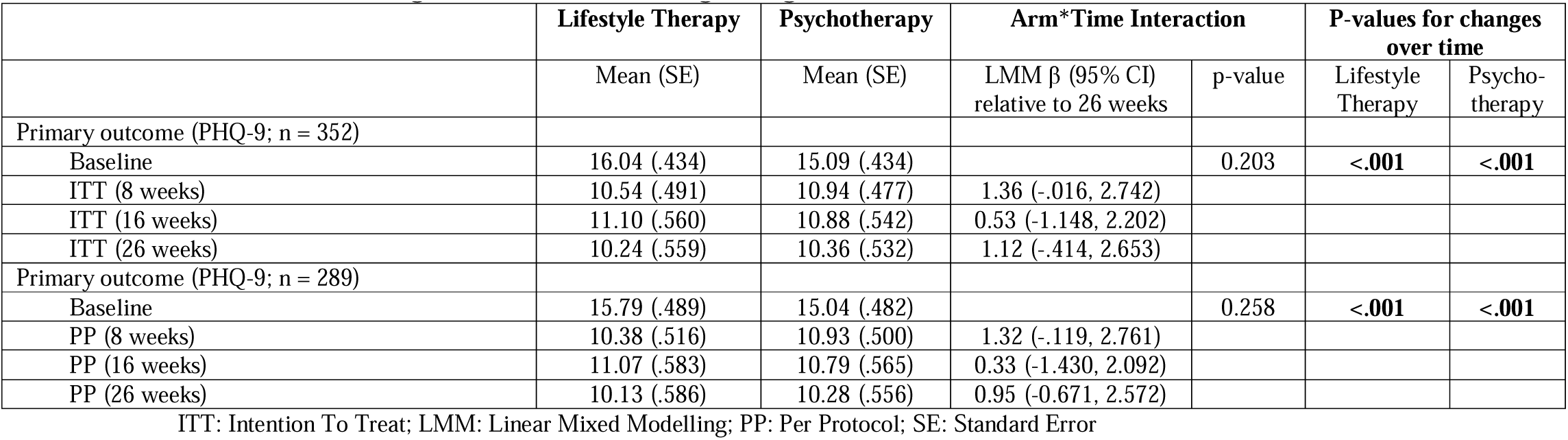
PHQ-9 Outcome using LMM and controlling for age.

|  | Lifestyle Therapy | Psychotherapy | Arm*Time Interaction |  | P-values for changes over time |  |
| --- | --- | --- | --- | --- | --- | --- |
| | Mean (SE) | Mean (SE) | LMM $\beta$ (95% CI)<br>relative to 26 weeks | p-value | Lifestyle<br>Therapy | Psycho-<br>therapy |
| Primary outcome (PHQ-9; n = 352) |  |  |  |  |  |  |
| Baseline | 16.04 (.434) | 15.09 (.434) |  | 0.203 | **<.001** | **<.001** |
| ITT (8 weeks) | 10.54 (.491) | 10.94 (.477) | 1.36 (-.016, 2.742) |  |  |  |
| ITT (16 weeks) | 11.10 (.560) | 10.88 (.542) | 0.53 (-1.148, 2.202) |  |  |  |
| ITT (26 weeks) | 10.24 (.559) | 10.36 (.532) | 1.12 (-.414, 2.653) |  |  |  |
| Primary outcome (PHQ-9; n = 289) |  |  |  |  |  |  |
| Baseline | 15.79 (.489) | 15.04 (.482) |  | 0.258 | **<.001** | **<.001** |
| PP (8 weeks) | 10.38 (.516) | 10.93 (.500) | 1.32 (-.119, 2.761) |  |  |  |
| PP (16 weeks) | 11.07 (.583) | 10.79 (.565) | 0.33 (-1.430, 2.092) |  |  |  |
| PP (26 weeks) | 10.13 (.586) | 10.28 (.556) | 0.95 (-0.671, 2.572) |  |  |  |

ITT: Intention To Treat; LMM: Linear Mixed Modelling; PP: Per Protocol; SE: Standard Error

Moderation analyses examined whether baseline imbalances influenced treatment response. No moderation effects were observed for personality disorder symptoms (p=0.620), self-efficacy (p=0.382), treatment expectancy (p=0.241) or medication adherence (p=0.139). However, readiness to change significantly moderated outcomes (p=0.024). Within lifestyle therapy, each one-unit increase in standardised readiness-to-change scores was associated with a 2.41-point reduction in MADRS at week 8 (95% CI: 0.60–4.22). No comparable effect was observed for psychotherapy.

Non-inferiority was examined separately in participants with MDD and BD. For MDD (n=291), the hypothesis was supported, as the 95% CI for the between-group difference in change scores (-0.34 to 4.06) remained above the non-inferiority margin of −1.6 MADRS points. For BD (n=67), results were inconclusive. While the observed effect was in the direction of lifestyle therapy, the CI (-3.86 to 7.13) was wide and included values indicating that lifestyle therapy could be clinically worse than CBT by more than the prespecified non-inferiority margin. Findings were consistent in per-protocol analyses. Treatment gains were maintained over 26 weeks, with non-inferiority on PHQ-9 change scores supported at 8, 16, and 26 weeks in both intention-to-treat (Supplementary Figure 1) and per-protocol analyses (Table 3).

### Secondary outcomes

No significant arm-by-assessment interactions were identified for secondary outcomes in either ITT or PP analyses, indicating similar changes over time between arms. Significant improvements across several outcomes were observed in both arms (Table 3). Lifestyle therapy participants also showed improvements in sleep, vegetable intake, alcohol and substance use risk, and self-efficacy. Participants in the psychotherapy arm demonstrated improvements in perceived social support.

Within the lifestyle therapy arm, 54.4% of participants improved dietary adherence by >15%, whereas 34.9% showed deterioration. Among those at moderate-to-high substance risk, 55.6% moved to a lower ASSIST risk category for alcohol, and 70.4% moved to a lower category for at least one substance. Clinical insomnia improved in 30.4% of participants, although 12.5% developed insomnia by week 8. For physical activity, 23.9% of those not meeting guidelines at baseline achieved recommended levels by week 8, while 31.2% who initially met guidelines no longer did so at week 8. In the psychotherapy arm, 35% achieved predefined CBT targets, with 35–42% meeting subscale targets, although 17.5–30.8% deteriorated.

### Safety and protocol deviations

Seven-hundred nineteen treatment-emergent safety events were recorded, including 687 adverse events, 17 adverse reactions, and 15 serious adverse events. Most events were mild or moderate and judged unrelated or unlikely related to the intervention (Supplementary Tables S4-6). At least one event was reported by 77.1% of participants in lifestyle therapy and 80.4% in psychotherapy. Protocol deviations (n=1149) were mainly missed sessions and incomplete assessments not affecting trial outcomes (Supplementary Table S7). Risk mitigation actions that occurred during unblinding included reassigning unblinded assessors.

## DISCUSSION

In this randomised non-inferiority trial involving adults with moderate-to-severe mood disorders, a multicomponent lifestyle intervention delivered by accredited dietitians and exercise physiologists achieved reductions in depressive symptoms that were non-inferior to a brief, structured cognitive behavioural therapy (CBT) programme delivered by psychologists. These findings provide evidence that lifestyle therapy can achieve comparable short-term clinical outcomes while being delivered by a broader allied health workforce. Given persistent international shortages of mental health professionals and growing demand for depression treatment, these findings suggest that appropriately trained allied health professionals could safely expand the workforce able to deliver evidence-based depression care.

Lifestyle therapies are increasingly recommended in international clinical guidelines, yet implementation in routine practice has been limited, in part because few studies have compared these interventions directly with established psychological treatments. Our findings, together with previous trials^10^ ^11^, strengthen the evidence supporting lifestyle therapy as an additional evidence-based treatment option for adults with mood disorders. Many health systems already employ dietitians and exercise professionals, and these workforces continue to expand internationally. Integrating these professions into collaborative mental healthcare could increase access to evidence-based depression treatment without relying solely on specialist mental health clinicians.

The magnitude of symptom improvement observed in both treatment groups was consistent with that reported in trials of brief psychological interventions for depression^15^, suggesting that lifestyle therapy achieved clinically meaningful improvements while maintaining comparable effectiveness to an established evidence-based treatment. Beyond reducing depressive symptoms, lifestyle therapy improved diet quality, sleep, and alcohol and substance use risk, suggesting that benefits may extend beyond mental health alone. These broader health gains are particularly relevant because people living with mood disorders experience substantially higher rates of cardiometabolic disease and premature mortality.^16^ Consistent with our previous trials^17^, participants receiving lifestyle therapy improved diet quality despite little overall change in physical activity. Improvements occurred independently of weight loss, supporting evidence that dietary quality itself may be an important therapeutic target for depression. The intervention therefore emphasised sustainable improvements in health behaviours rather than weight reduction.

The findings should not be interpreted as suggesting that lifestyle therapy replaces psychological treatment. Rather, lifestyle interventions may expand the range of evidence-based options available to clinicians and patients. Depending on individual preferences, clinical formulation, and service availability, lifestyle therapy could be delivered as an initial treatment option, integrated with psychotherapy, or incorporated into maintenance and relapse prevention strategies. These findings therefore support current international guideline recommendations that lifestyle interventions should form a foundational component of comprehensive mood disorder care^6^. Indeed, these interventions could be incorporated into collaborative care or stepped-care models already operating in many health systems, enabling psychologists to focus on patients requiring higher-intensity psychological treatment. Lifestyle therapy may also appeal to people who prefer behavioural approaches or who are unwilling or unable to engage in psychological therapy, supporting greater choice within evidence-based depression care.

### Strengths

This is the largest randomised trial to compare a multicomponent lifestyle intervention with a structured CBT programme for adults with moderate-to-severe mood disorders. The study addresses important limitations of previous lifestyle psychiatry trials^10^ ^11^ through diagnostic confirmation of mood disorders, interviewer-rated primary outcomes, a prospectively registered non-inferiority design, and comparison with an active psychological treatment rather than usual care or an attention control. Both interventions were manualised, delivered with high treatment fidelity by appropriately qualified clinicians, and matched for treatment dose, clinician contact, delivery format, and group structure. Participant engagement was high, with more than 80% receiving the predefined therapeutic dose, and treatment effects were maintained over six months. The telehealth model, together with prospective monitoring of safety outcomes, further supports the feasibility and scalability of this approach within routine healthcare.

### Limitations

Several limitations should be considered. Participants were predominantly White, highly educated women, reflecting the demographic profile commonly observed in lifestyle^10^ ^11^ and psychological treatment trials^17^, and potentially limiting generalisability to more diverse populations. Although recruitment achieved 95% of the planned sample size and the primary findings were consistent across intention-to-treat, per-protocol, and sensitivity analyses, the bipolar disorder subgroup was comparatively small and conclusions for this population remain uncertain. As in most behavioural intervention trials, participants and clinicians could not be blinded to treatment allocation, although outcome assessors remained masked and interviewer-rated outcomes reduced the potential for assessment bias. Finally, the comparator was a brief, structured group CBT programme delivered under pragmatic conditions. The findings therefore support non-inferiority relative to this treatment model and should not be interpreted as demonstrating equivalence with all CBT formats or treatment intensities.

## CONCLUSION

A multicomponent lifestyle intervention delivered by accredited dietitians and exercise physiologists was non-inferior to a brief structured CBT programme for adults with moderate-to-severe mood disorders. These findings support lifestyle therapy as an additional evidence-based treatment option that could expand access to effective depression care by broadening the clinical workforce able to deliver evidence-based depression care. As health systems seek scalable responses to growing demand for mental healthcare, implementation and cost-effectiveness studies should now determine how best to integrate lifestyle therapy into routine clinical practice.

## Data Availability

All data produced in the present study are available upon reasonable request to the authors

https://researchdata.edu.au/health/

## Study contributors

*Study conceptualisation:* AO, FNJ, MBe *Trial enactment & oversight:* AO, KR, JAD, TJo, SM, MBr, RF, SB, DS, MLC, MT, FNJ, NLM, MG, LKR, EMCD, MC, GM, NT, MO, TR, ESG, WM, TJa, AR, MB. *Manuscript preparation:* AO, KR, DM, MG. *Manuscript revision:* AO, KR, JAD, TJo, SM, MBr, RF, DS, MLC, MT, FNJ, NLM, MG, LKR, EMCD, MC, GM, NT, MO, TR, ESG, WM, TJa, AR, DM, PM, RI, MBe. All authors read, approved and take responsibility for the final manuscript. AO takes final responsibility for the decision to submit the manuscript.

## Data Sharing Statement

A minimum dataset of individual participant data will be made available via https://researchdata.edu.au/health/ after de-identification for any purpose approved by the applicant’s Human Research Ethics Committee and the HARMON-E Chief Investigator Team. This will be available immediately following publication with no end date.

## AI use

During the preparation of this work, the author(s) used ChatGPT for light editing purposes. The author(s) reviewed and edited the output as needed and take full responsibility for the content of the published article.

## Acknowledgements

DM, PM and KR verified the dataset and have access to the raw data. The authors would like to acknowledge the contribution of our DSMB members – Prof Philip Boyce, Prof Jane Speight, Dr Stella May Gwini, and Dr Debbie Ashtree. We would also like to thank our participants who volunteered their time for this study and all project team members.

## Abbreviations

CBT: Cognitive Behavioural Therapy
MADRS: Montgomery–Åsberg Depression Rating Scale
LMM: Linear Mixed Model
CI: Confidence Interval
ITT: Intention-to-Treat
PP: Per-Protocol
MDD: Major Depressive Disorder
BD: Bipolar Disorder
CONSORT: Consolidated Standards of Reporting Trials
HREC: Human Research Ethics Committee
ICH: International Council for Harmonisation
SCID-5 RV: Structured Clinical Interview for DSM-5 Research Version
REDCap: Research Electronic Data Capture
CALM: Curbing Anxiety and Depression using Lifestyle Medicine
PHQ-9: Patient Health Questionnaire-9
ASSIST: Alcohol Smoking and Substance Involvement Screening Test
ISI: Insomnia Severity Index
SIMPAQ: Simple Physical Activity Questionnaire
CBT-SUITS: Cognitive Behavioural Therapy Suitability Scale
DASS-10: Depression Anxiety and Stress Scale
DSMB: Data Safety Monitoring Board
ICC: Intraclass Correlation Coefficient
ANCOVA: Analysis of Covariance
BDRS: Bipolar Depression Rating Scale
GAD-7: Generalized Anxiety Disorder-7
MHRM-10: Mental Health Recovery Measure-10
MOS-SSS: Medical Outcomes Study Social Support Scale
GSE: General Self-Efficacy Scale
AQoL-4D: Assessment of Quality of Life–4 Dimension Utility Instrument
BMI: Body Mass Index.

## SUPPLEMENTARY MATERIALS

**Supplementary Figure 1.**
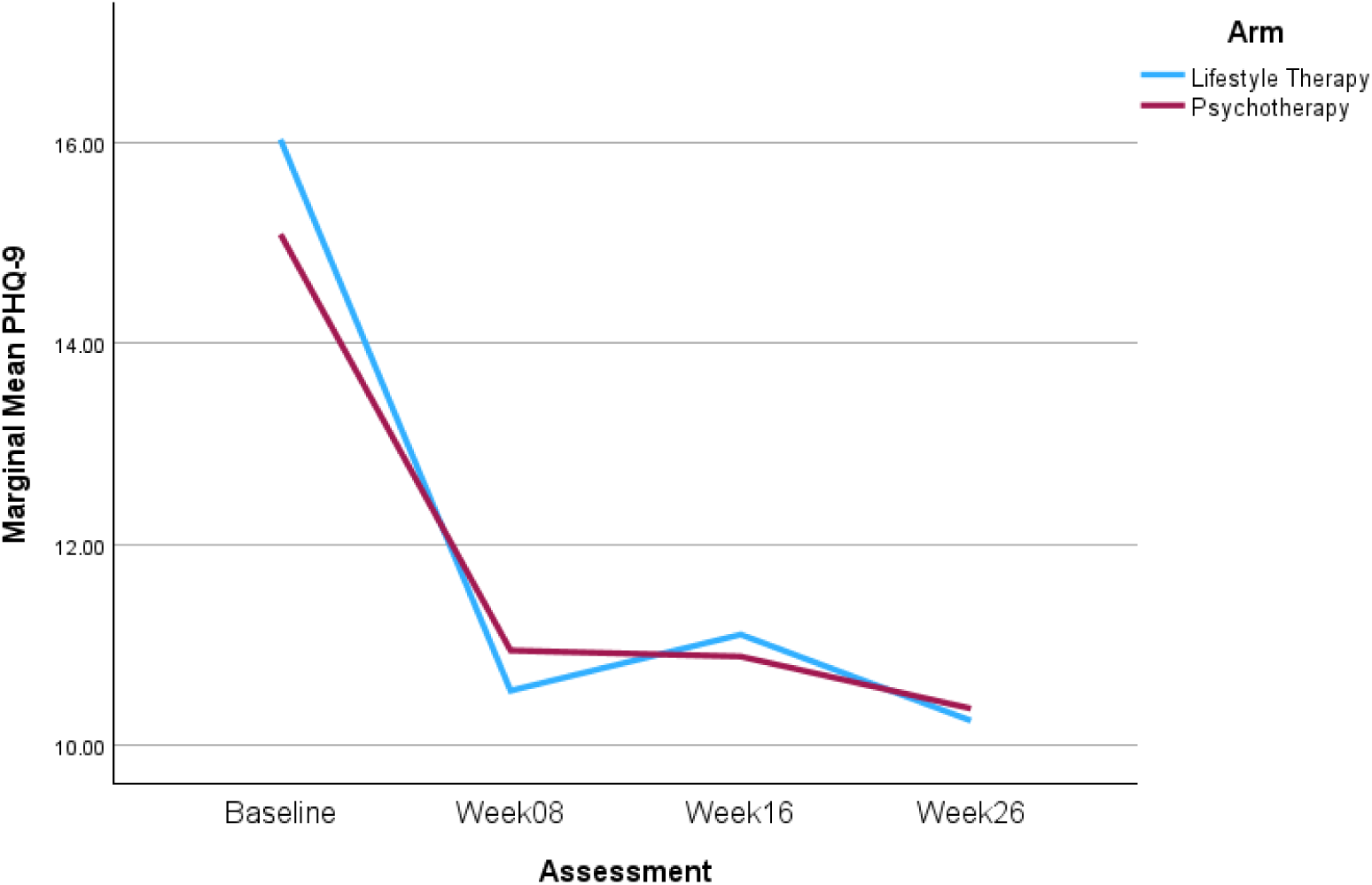
Intention To Treat analysis for PHQ-9 depression over 26 weeks controlling for age at enrolment

**Supplementary Figure 2.**
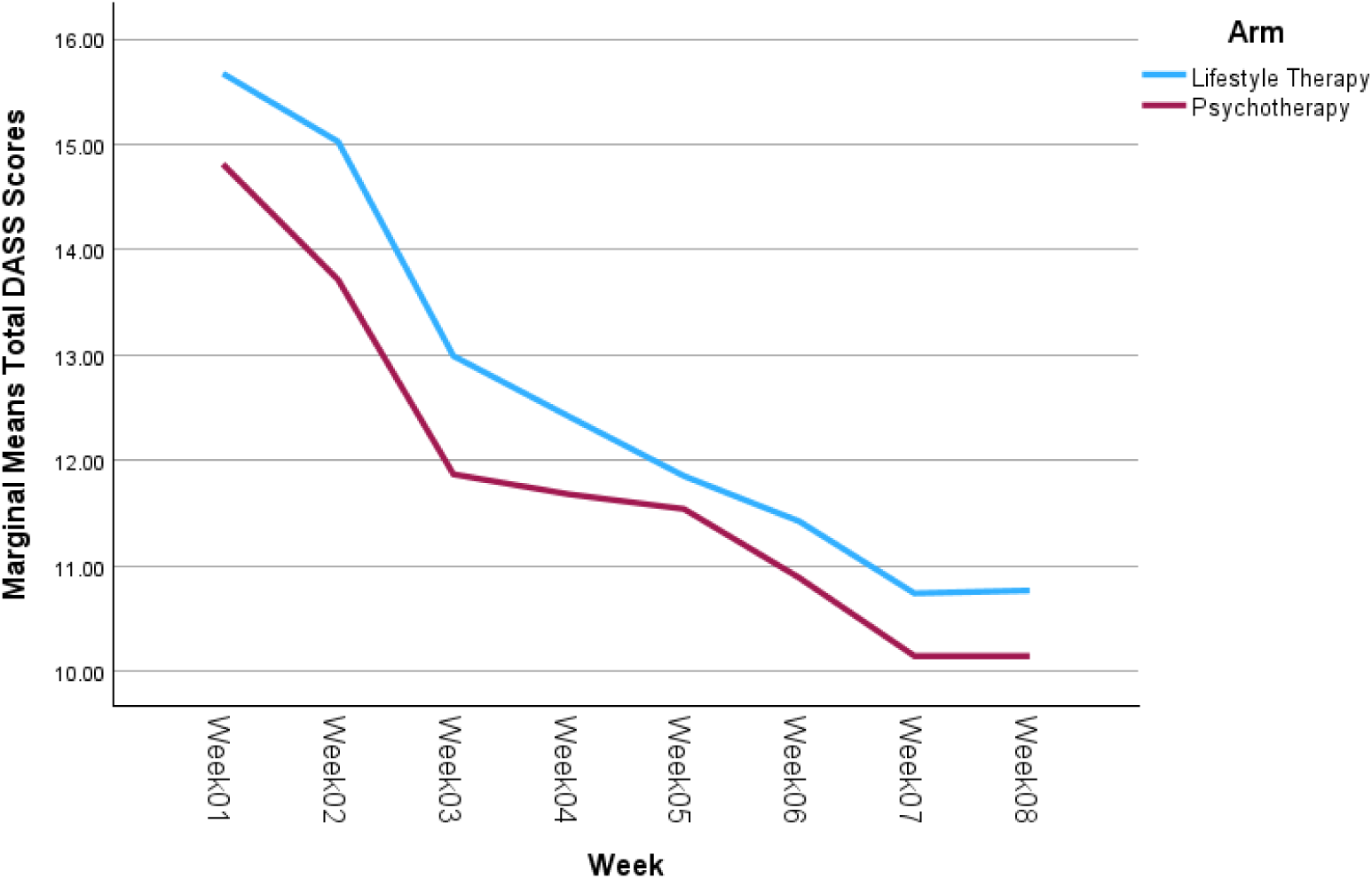
Intention To Treat analysis for Distress Anxiety and Stress Scales psychological distress (safety scores) over 8 weeks

**Table S1.**
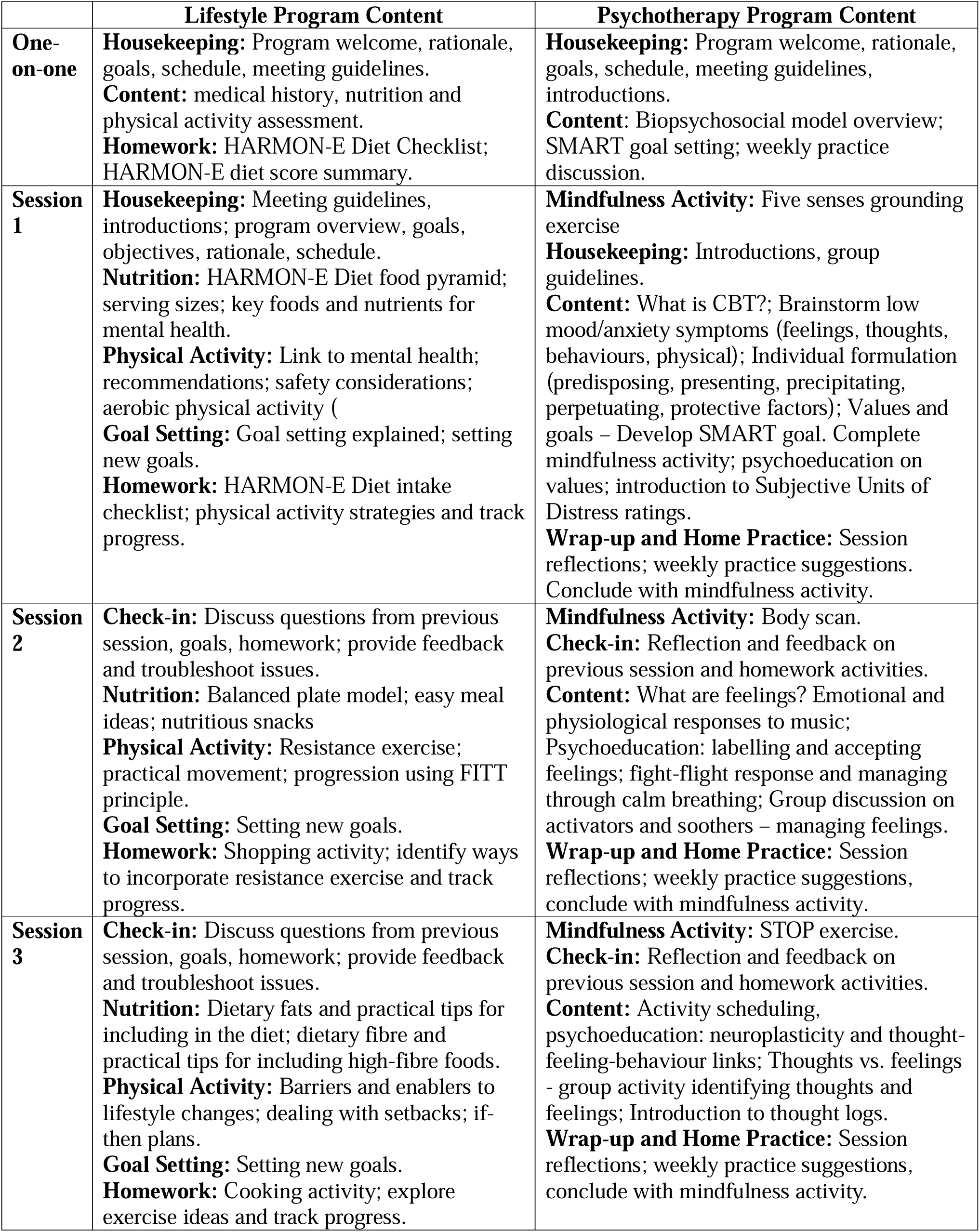
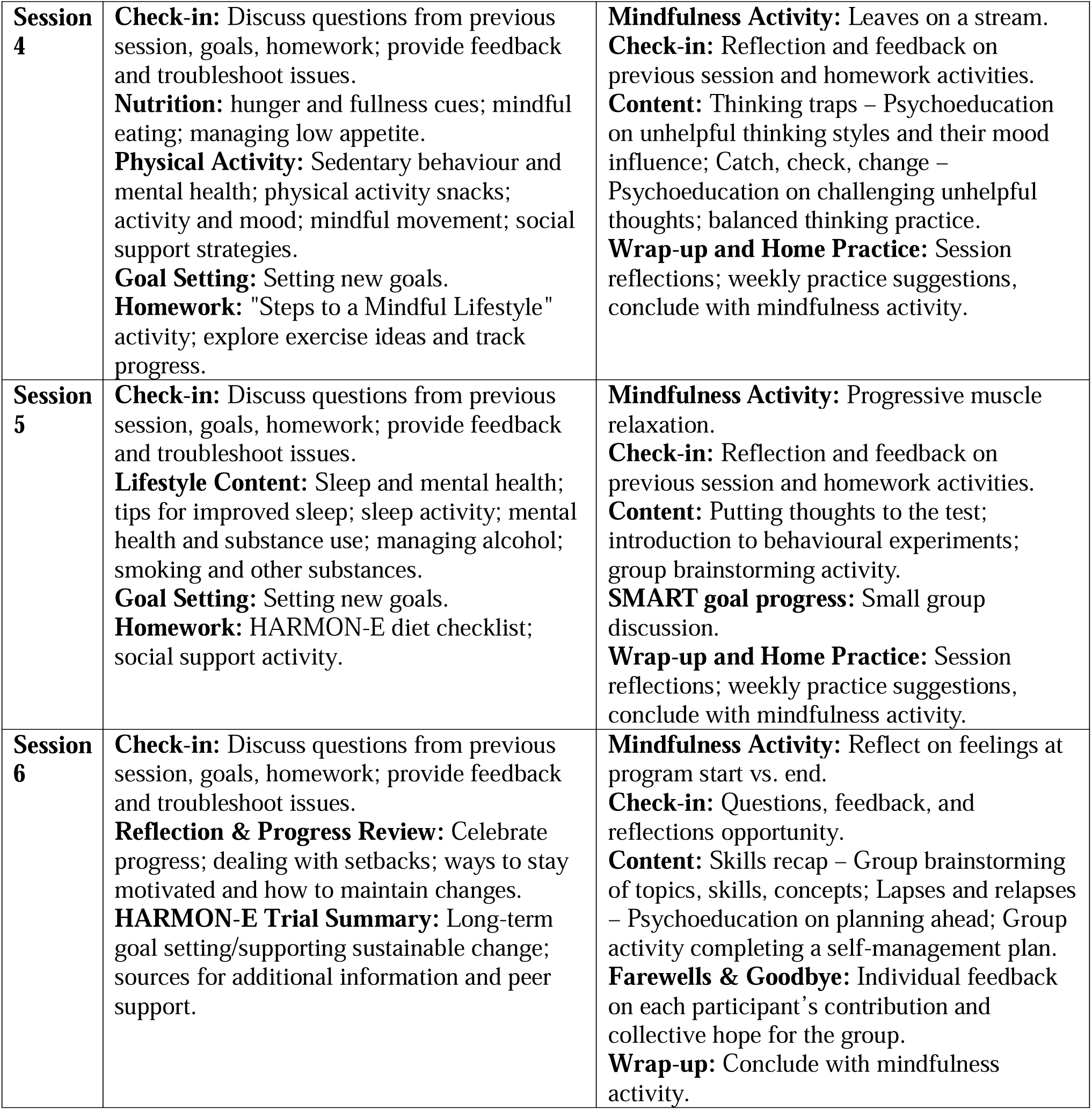
Intervention content overview.

**Table S2.**
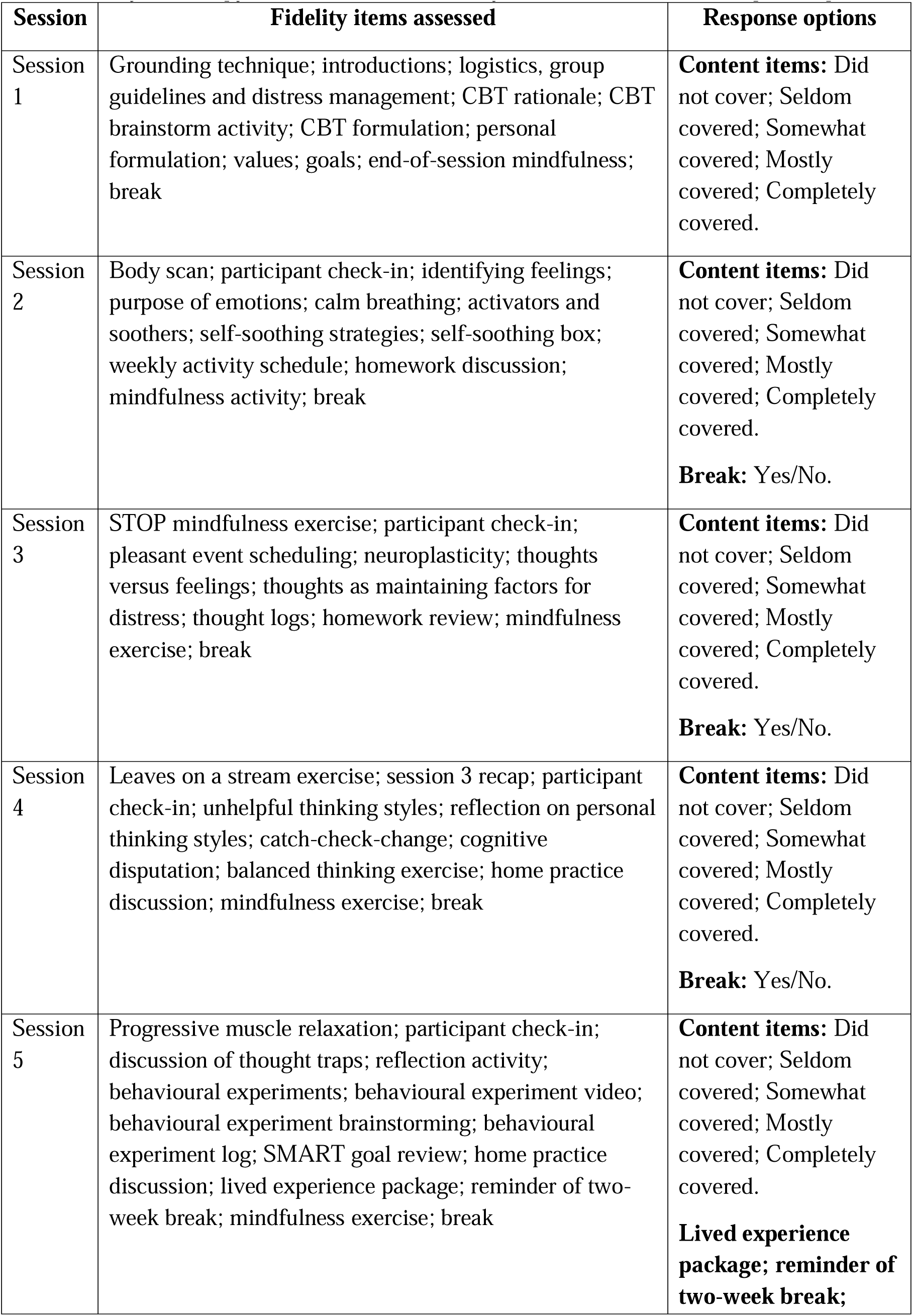
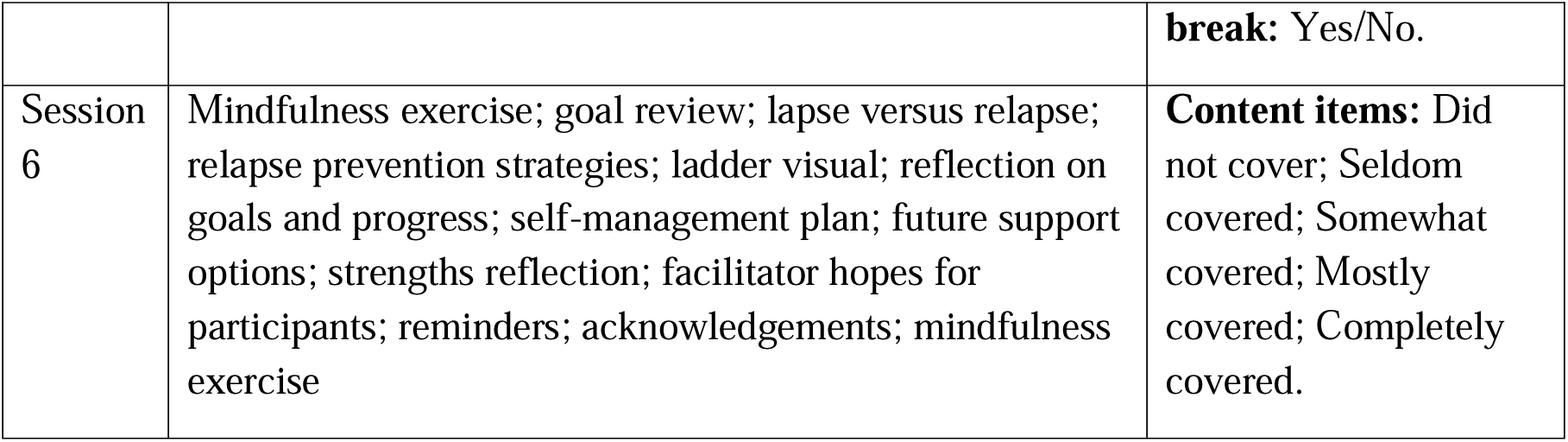
Psychotherapy (CBT) intervention fidelity checklist domains and response options.

**Table S3.**
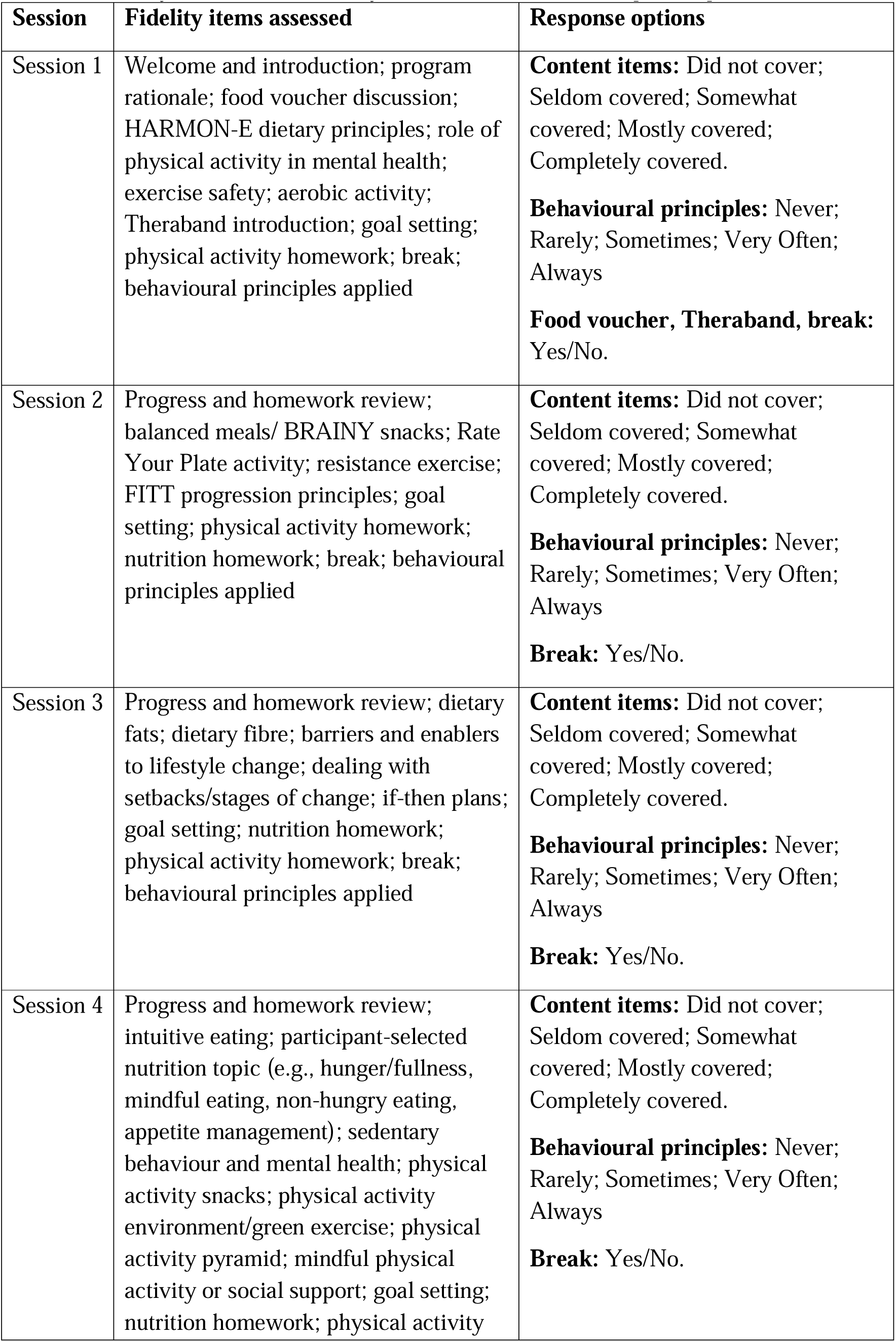
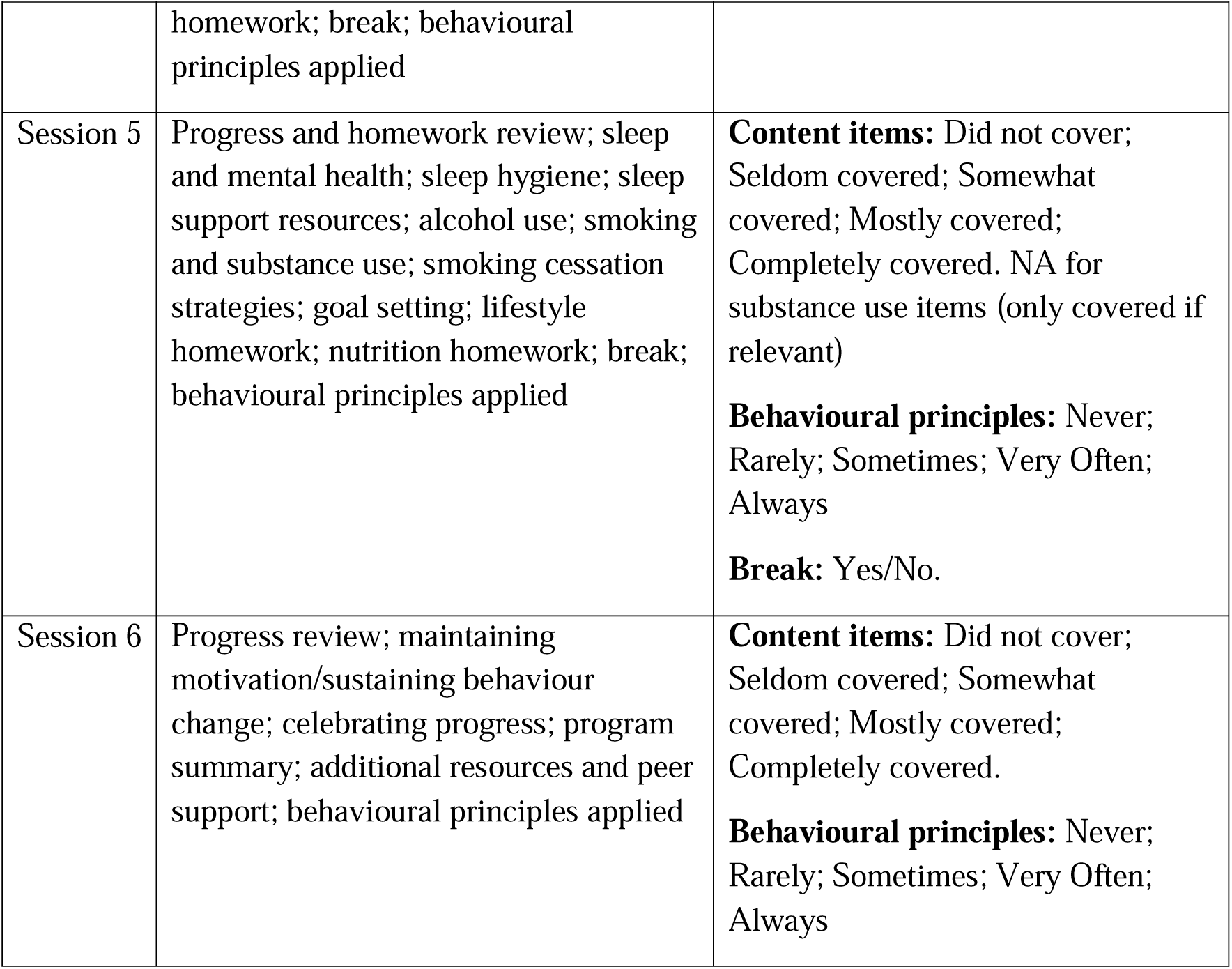
Lifestyle intervention fidelity checklist domains and response options.

**Table S4.**
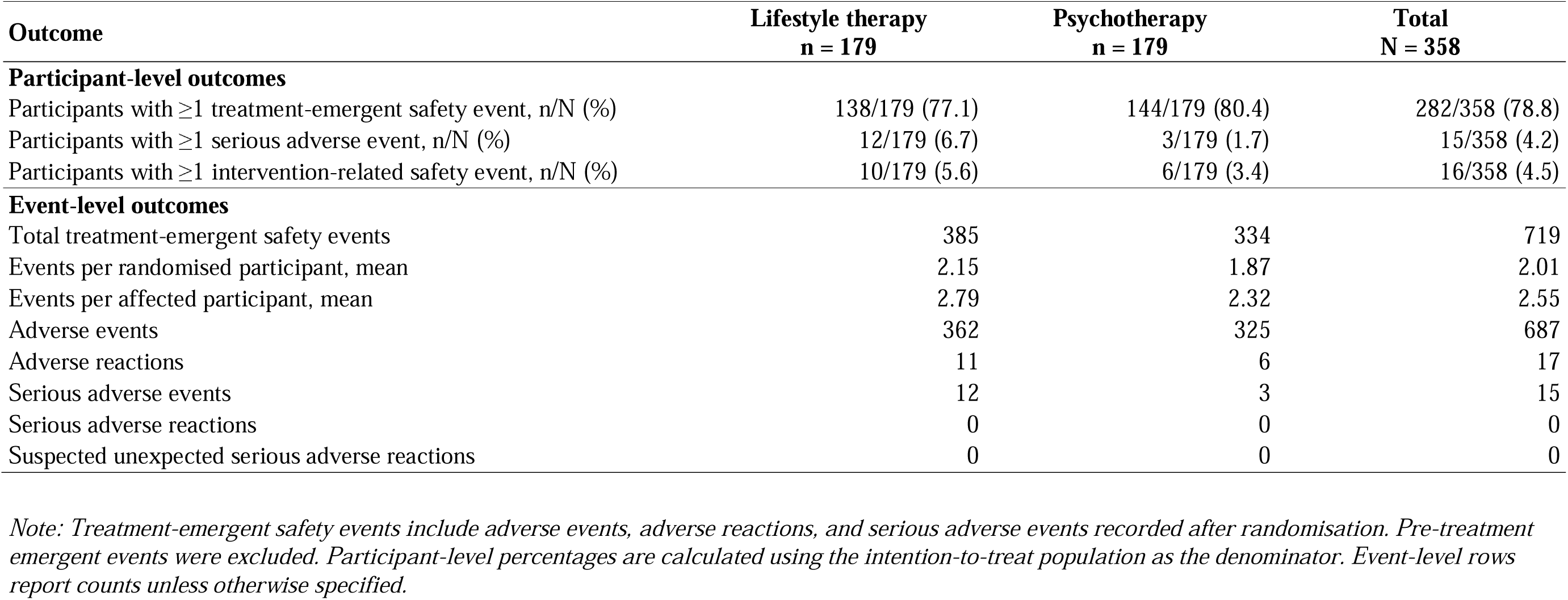
Summary of treatment-emergent safety events by randomised group, intention-to-treat population.

**Table S5.**
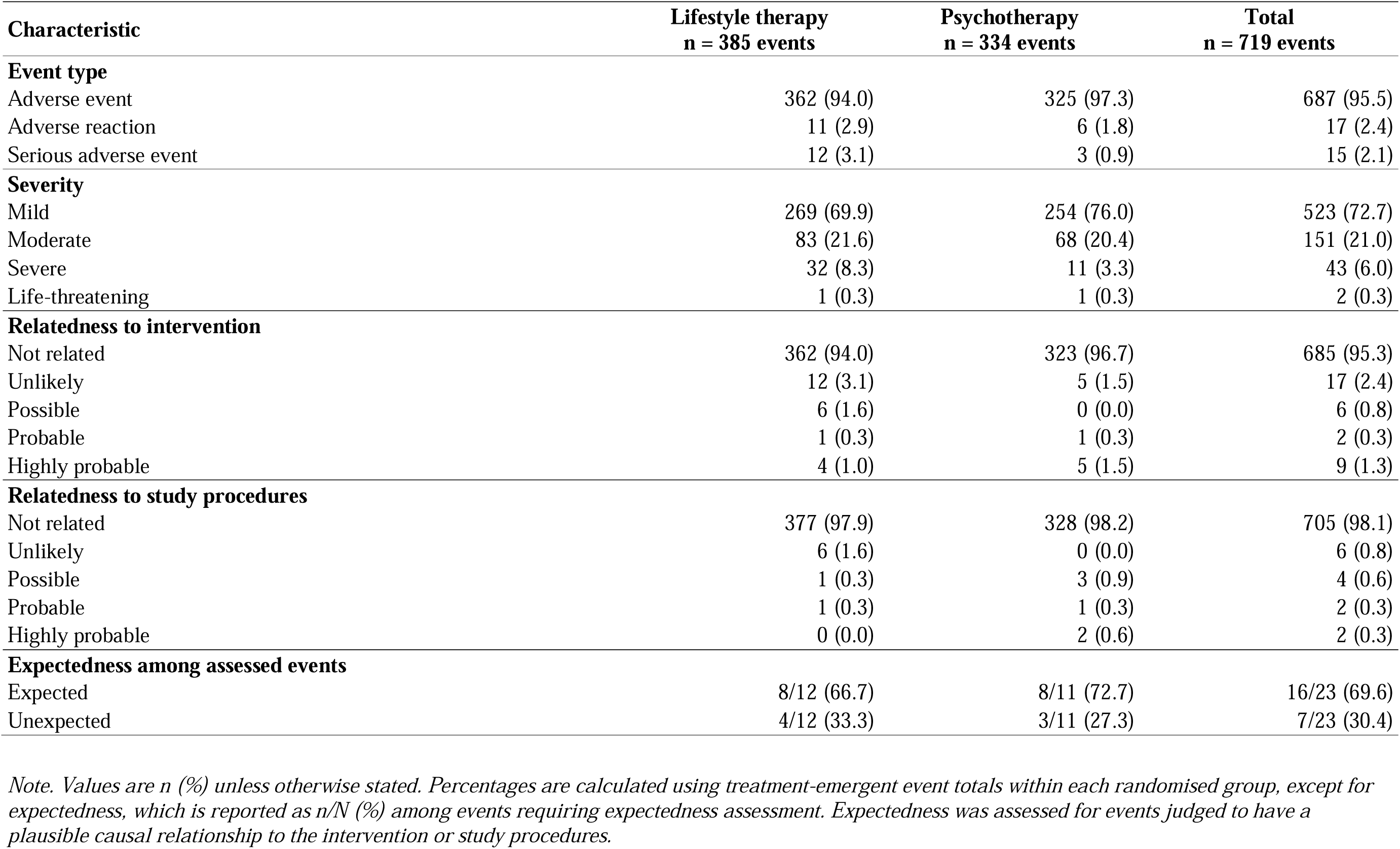
Classification and adjudication characteristics of treatment-emergent safety events by randomised group, intention-to-treat population.

**Table S6.**
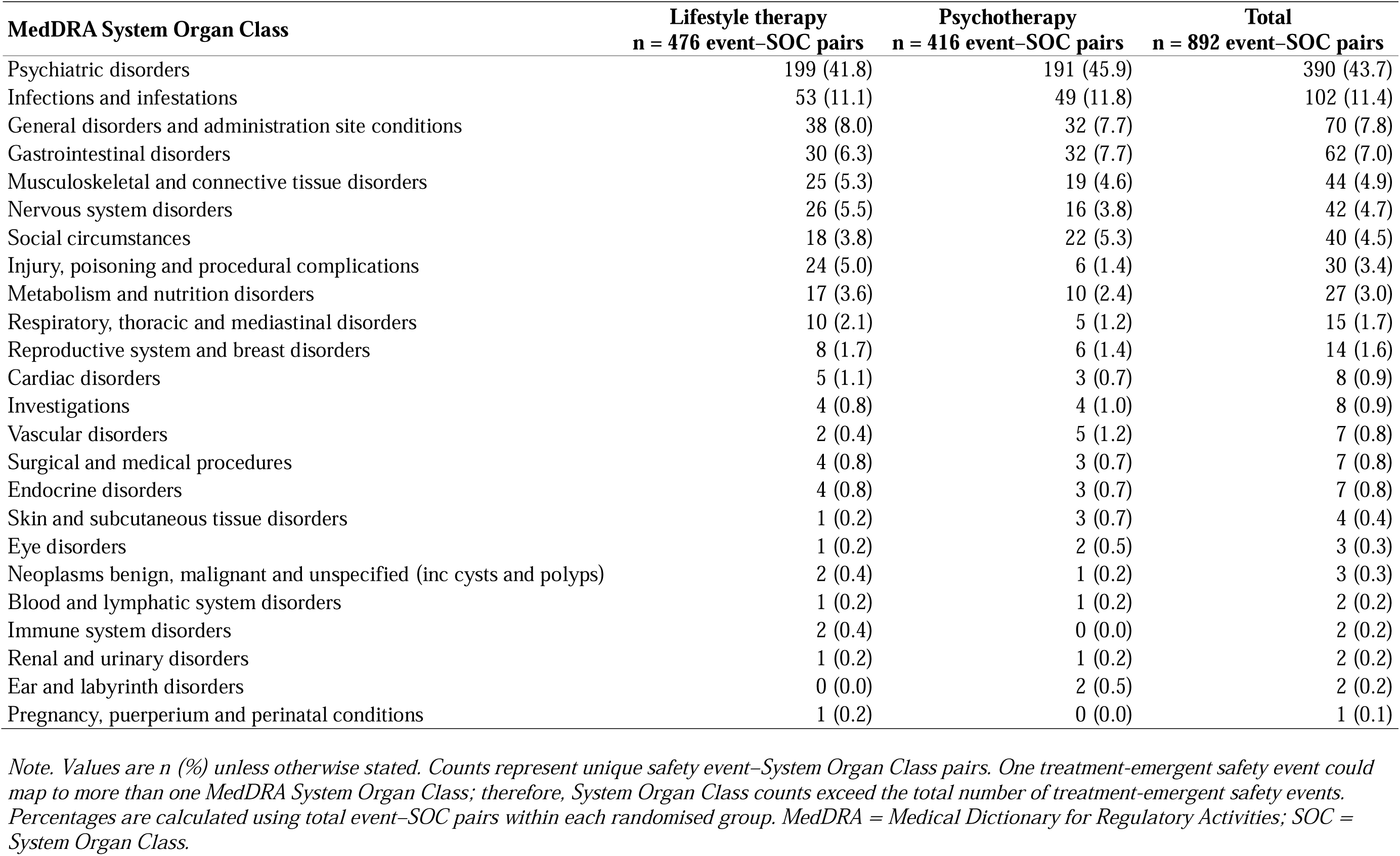
Treatment-emergent safety event–System Organ Class pairs by randomised group, intention-to-treat population.

**Table S7.**
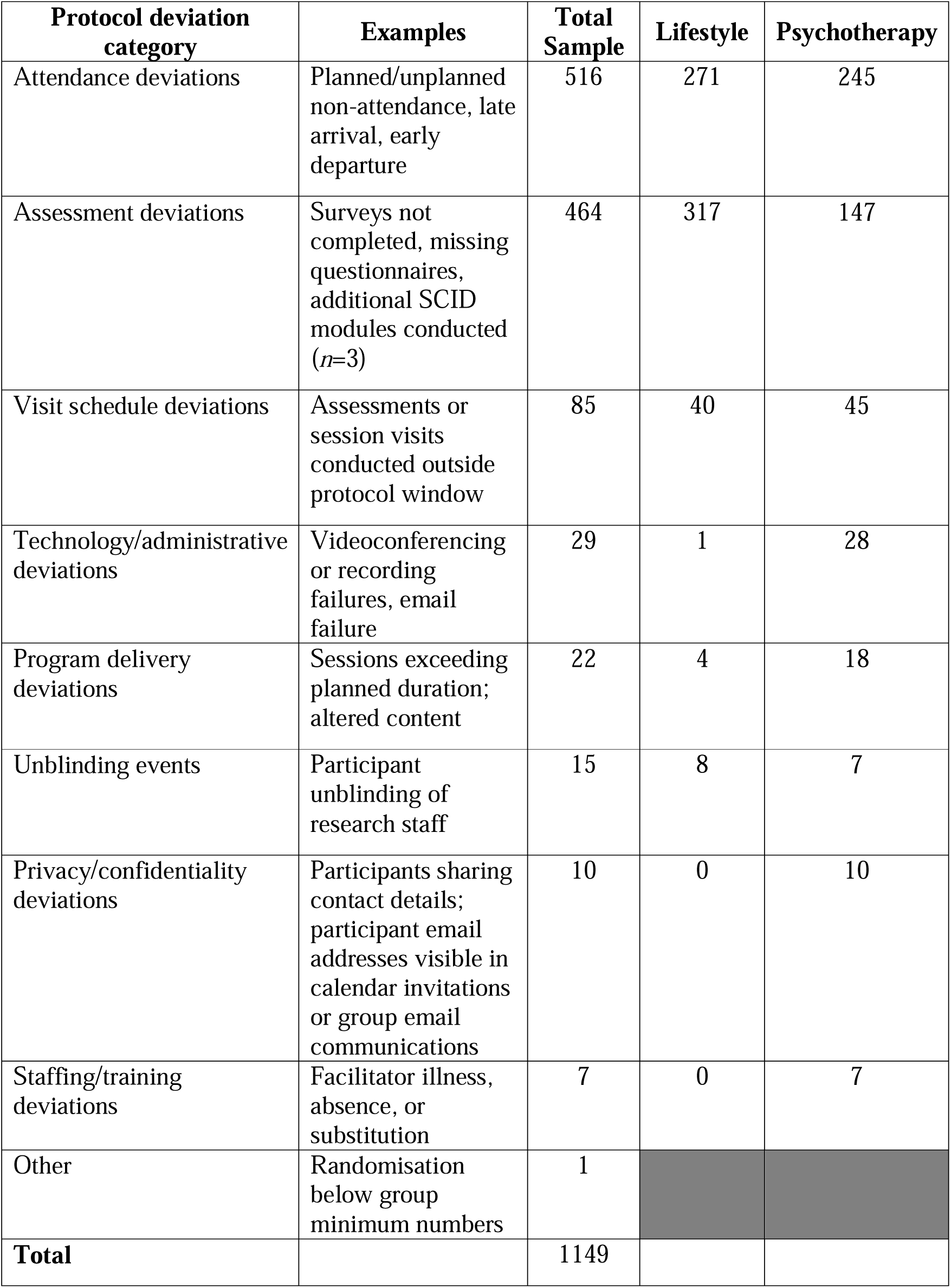
Protocol deviations for total sample and intervention arms.

