## Supplementary Figure 1 for "Lifestyle therapy versus cognitive behavioural therapy for adults with mood disorders: a randomised non-inferiority trial"

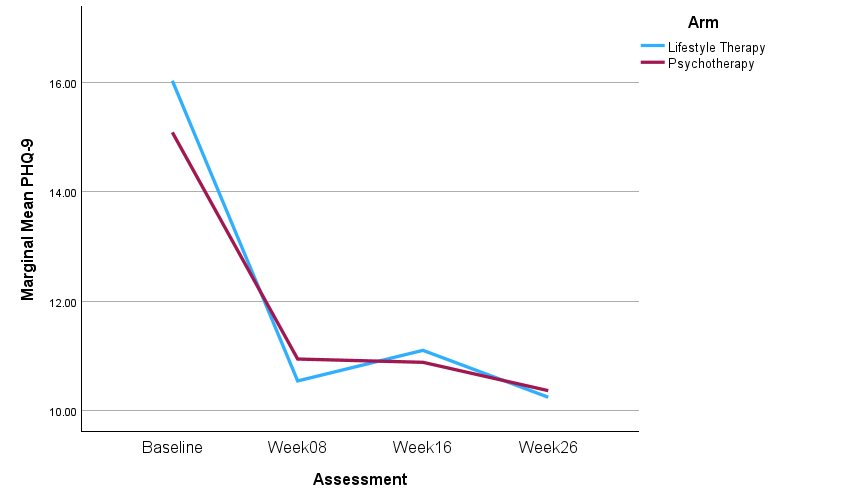

*Figure 3. Intention To Treat analysis for PHQ-9 depression over 26 weeks controlling for age at enrolment*
