## Supplementary Figure 2 for "Lifestyle therapy versus cognitive behavioural therapy for adults with mood disorders: a randomised non-inferiority trial"

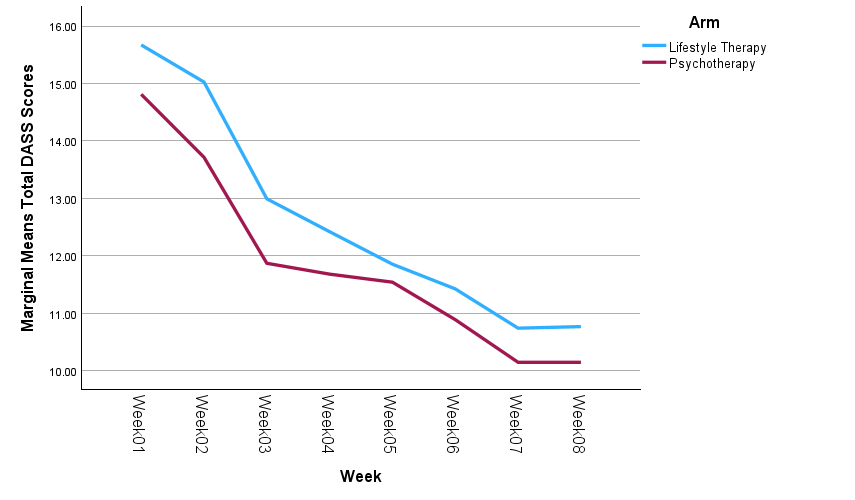

*Figure 4.* *Intention To Treat analysis for Distress Anxiety and Stress Scales psychological distress (safety scores) over 8-week*
