## Supplementary Table 1 for "Lifestyle therapy versus cognitive behavioural therapy for adults with mood disorders: a randomised non-inferiority trial"

*Table 1. Intervention content overview*

|  | **Lifestyle Program Content** | **Psychotherapy Program Content** |
| --- | --- | --- |
| **One-on-one** | **Housekeeping:** Program welcome, rationale, goals, schedule, meeting guidelines.  **Content:** medical history, nutrition and physical activity assessment.  **Homework:** HARMON-E Diet Checklist; HARMON-E diet score summary. | **Housekeeping:** Program welcome, rationale, goals, schedule, meeting guidelines, introductions.  **Content:** Biopsychosocial model overview; SMART goal setting; weekly practice discussion. |
| **Session 1** | **Housekeeping:** Meeting guidelines, introductions; program overview, goals, objectives, rationale, schedule.  **Nutrition:** HARMON-E Diet food pyramid; serving sizes; key foods and nutrients for mental health.  **Physical Activity:** Link to mental health; recommendations; safety considerations; aerobic physical activity (  **Goal Setting:** Goal setting explained; setting new goals.  **Homework:** HARMON-E Diet intake checklist; physical activity strategies and track progress. | **Mindfulness Activity: Five senses grounding exercise**  **Housekeeping:** Introductions, group guidelines.  **Content:** What is CBT?; Brainstorm low mood/anxiety symptoms (feelings, thoughts, behaviours, physical); Individual formulation (predisposing, presenting, precipitating, perpetuating, protective factors); Values and goals – Develop SMART goal. Complete mindfulness activity; psychoeducation on values; introduction to Subjective Units of Distress ratings.  **Wrap-up and Home Practice:** Session reflections; weekly practice suggestions. Conclude with mindfulness activity. |
| **Session 2** | **Check-in:** Discuss questions from previous session, goals, homework; provide feedback and troubleshoot issues.  **Nutrition:** Balanced plate model; easy meal ideas; nutritious snacks  **Physical Activity:** Resistance exercise; practical movement; progression using FITT principle.  **Goal Setting:** Setting new goals.  **Homework:** Shopping activity; identify ways to incorporate resistance exercise and track progress. | **Mindfulness Activity:** Body scan.  **Check-in:** Reflection and feedback on previous session and homework activities.  **Content:** What are feelings? Emotional and physiological responses to music; Psychoeducation: labelling and accepting feelings; fight-flight response and managing through calm breathing; Group discussion on activators and soothers – managing feelings.  **Wrap-up and Home Practice:** Session reflections; weekly practice suggestions, conclude with mindfulness activity. |
| **Session 3** | **Check-in:** Discuss questions from previous session, goals, homework; provide feedback and troubleshoot issues.  **Nutrition:** Dietary fats and practical tips for including in the diet; dietary fibre and practical tips for including high-fibre foods.  **Physical Activity:** Barriers and enablers to lifestyle changes; dealing with setbacks; if-then plans.  **Goal Setting:** Setting new goals.  **Homework:** Cooking activity; explore exercise ideas and track progress. | **Mindfulness Activity:** STOP exercise.  **Check-in:** Reflection and feedback on previous session and homework activities.  **Content:** Activity scheduling, psychoeducation: neuroplasticity and thought-feeling-behaviour links; Thoughts vs. feelings - group activity identifying thoughts and feelings; Introduction to thought logs.  **Wrap-up and Home Practice:** Session reflections; weekly practice suggestions, conclude with mindfulness activity. |
| **Session 4** | **Check-in:** Discuss questions from previous session, goals, homework; provide feedback and troubleshoot issues.  **Nutrition:** hunger and fullness cues; mindful eating; managing low appetite.  **Physical Activity:** Sedentary behaviour and mental health; physical activity snacks; activity and mood; mindful movement; social support strategies.  **Goal Setting:** Setting new goals.  **Homework:** "Steps to a Mindful Lifestyle" activity; explore exercise ideas and track progress. | **Mindfulness Activity:** Leaves on a stream.  **Check-in:** Reflection and feedback on previous session and homework activities.  **Content:** Thinking traps – Psychoeducation on unhelpful thinking styles and their mood influence; Catch, check, change – Psychoeducation on challenging unhelpful thoughts; balanced thinking practice.  **Wrap-up and Home Practice:** Session reflections; weekly practice suggestions, conclude with mindfulness activity. |
| **Session 5** | **Check-in:** Discuss questions from previous session, goals, homework; provide feedback and troubleshoot issues.  **Lifestyle Content:** Sleep and mental health; tips for improved sleep; sleep activity; mental health and substance use; managing alcohol; smoking and other substances.  **Goal Setting:** Setting new goals.  **Homework:** HARMON-E diet checklist; social support activity. | **Mindfulness Activity:** Progressive muscle relaxation.  **Check-in:** Reflection and feedback on previous session and homework activities.  **Content:** Putting thoughts to the test; introduction to behavioural experiments; group brainstorming activity.  **SMART goal progress:** Small group discussion.  **Wrap-up and Home Practice:** Session reflections; weekly practice suggestions, conclude with mindfulness activity. |
| **Session 6** | **Check-in:** Discuss questions from previous session, goals, homework; provide feedback and troubleshoot issues.  **Reflection & Progress Review:** Celebrate progress; dealing with setbacks; ways to stay motivated and how to maintain changes.  **HARMON-E Trial Summary:** Long-term goal setting/supporting sustainable change; sources for additional information and peer support. | **Mindfulness Activity: Reflect on feelings at program start vs. end.**  **Check-in: Questions, feedback, and reflections opportunity.**  **Content: Skills recap – Group brainstorming of topics, skills, concepts; Lapses and relapses – Psychoeducation on planning ahead; Group activity completing a self-management plan. Farewells & Goodbye: Individual feedback on each participant’s contribution and collective hope for the group.**  **Wrap-up: Conclude with mindfulness activity.** |
