## Supplementary Table 2 for "Lifestyle therapy versus cognitive behavioural therapy for adults with mood disorders: a randomised non-inferiority trial"

Table S1. Psychotherapy (CBT) intervention fidelity checklist domains and response options

| **Session** | **Fidelity items assessed** | **Response options** |
| --- | --- | --- |
| Session 1 | Grounding technique; introductions; logistics, group guidelines and distress management; CBT rationale; CBT brainstorm activity; CBT formulation; personal formulation; values; goals; end-of-session mindfulness; break | **Content items:** Did not cover; Seldom covered; Somewhat covered; Mostly covered; Completely covered. |
| Session 2 | Body scan; participant check-in; identifying feelings; purpose of emotions; calm breathing; activators and soothers; self-soothing strategies; self-soothing box; weekly activity schedule; homework discussion; mindfulness activity; break | **Content items:** Did not cover; Seldom covered; Somewhat covered; Mostly covered; Completely covered.  **Break:** Yes/No. |
| Session 3 | STOP mindfulness exercise; participant check-in; pleasant event scheduling; neuroplasticity; thoughts versus feelings; thoughts as maintaining factors for distress; thought logs; homework review; mindfulness exercise; break | **Content items:** Did not cover; Seldom covered; Somewhat covered; Mostly covered; Completely covered.  **Break:** Yes/No. |
| Session 4 | Leaves on a stream exercise; session 3 recap; participant check-in; unhelpful thinking styles; reflection on personal thinking styles; catch-check-change; cognitive disputation; balanced thinking exercise; home practice discussion; mindfulness exercise; break | **Content items:** Did not cover; Seldom covered; Somewhat covered; Mostly covered; Completely covered.  **Break:** Yes/No. |
| Session 5 | Progressive muscle relaxation; participant check-in; discussion of thought traps; reflection activity; behavioural experiments; behavioural experiment video; behavioural experiment brainstorming; behavioural experiment log; SMART goal review; home practice discussion; lived experience package; reminder of two-week break; mindfulness exercise; break | **Content items:** Did not cover; Seldom covered; Somewhat covered; Mostly covered; Completely covered.  **Lived experience package; reminder of two-week break; break:** Yes/No. |
| Session 6 | Mindfulness exercise; goal review; lapse versus relapse; relapse prevention strategies; ladder visual; reflection on goals and progress; self-management plan; future support options; strengths reflection; facilitator hopes for participants; reminders; acknowledgements; mindfulness exercise | **Content items:** Did not cover; Seldom covered; Somewhat covered; Mostly covered; Completely covered. |
