## Supplementary Table 3 for "Lifestyle therapy versus cognitive behavioural therapy for adults with mood disorders: a randomised non-inferiority trial"

Table S2. Lifestyle intervention fidelity checklist domains and response options

| **Session** | **Fidelity items assessed** | **Response options** |
| --- | --- | --- |
| Session 1 | Welcome and introduction; program rationale; food voucher discussion; HARMON-E dietary principles; role of physical activity in mental health; exercise safety; aerobic activity; Theraband introduction; goal setting; physical activity homework; break; behavioural principles applied | **Content items:** Did not cover; Seldom covered; Somewhat covered; Mostly covered; Completely covered.  **Behavioural principles:** Never; Rarely; Sometimes; Very Often; Always  **Food voucher, Theraband, break:** Yes/No. |
| Session 2 | Progress and homework review; balanced meals/ BRAINY snacks; Rate Your Plate activity; resistance exercise; FITT progression principles; goal setting; physical activity homework; nutrition homework; break; behavioural principles applied | **Content items:** Did not cover; Seldom covered; Somewhat covered; Mostly covered; Completely covered.  **Behavioural principles:** Never; Rarely; Sometimes; Very Often; Always  **Break:** Yes/No. |
| Session 3 | Progress and homework review; dietary fats; dietary fibre; barriers and enablers to lifestyle change; dealing with setbacks/stages of change; if-then plans; goal setting; nutrition homework; physical activity homework; break; behavioural principles applied | **Content items:** Did not cover; Seldom covered; Somewhat covered; Mostly covered; Completely covered.  **Behavioural principles:** Never; Rarely; Sometimes; Very Often; Always  **Break:** Yes/No. |
| Session 4 | Progress and homework review; intuitive eating; participant-selected nutrition topic (e.g., hunger/fullness, mindful eating, non-hungry eating, appetite management); sedentary behaviour and mental health; physical activity snacks; physical activity environment/green exercise; physical activity pyramid; mindful physical activity or social support; goal setting; nutrition homework; physical activity homework; break; behavioural principles applied | **Content items:** Did not cover; Seldom covered; Somewhat covered; Mostly covered; Completely covered.  **Behavioural principles:** Never; Rarely; Sometimes; Very Often; Always  **Break:** Yes/No. |
| Session 5 | Progress and homework review; sleep and mental health; sleep hygiene; sleep support resources; alcohol use; smoking and substance use; smoking cessation strategies; goal setting; lifestyle homework; nutrition homework; break; behavioural principles applied | **Content items:** Did not cover; Seldom covered; Somewhat covered; Mostly covered; Completely covered. NA for substance use items (only covered if relevant)  **Behavioural principles:** Never; Rarely; Sometimes; Very Often; Always  **Break:** Yes/No. |
| Session 6 | Progress review; maintaining motivation/sustaining behaviour change; celebrating progress; program summary; additional resources and peer support; behavioural principles applied | **Content items:** Did not cover; Seldom covered; Somewhat covered; Mostly covered; Completely covered.  **Behavioural principles:** Never; Rarely; Sometimes; Very Often; Always |
