## Supplementary Table 4-6 for "Lifestyle therapy versus cognitive behavioural therapy for adults with mood disorders: a randomised non-inferiority trial"

*Table S3. Summary of treatment-emergent safety events by randomised group, intention-to-treat population*

| **Outcome** | **Lifestyle therapy**  **n = 179** | **Psychotherapy**  **n = 179** | **Total**  **N = 358** |
| --- | --- | --- | --- |
| **Participant-level outcomes** |  |  |  |
| Participants with ≥1 treatment-emergent safety event, n/N (%) | 138/179 (77.1) | 144/179 (80.4) | 282/358 (78.8) |
| Participants with ≥1 serious adverse event, n/N (%) | 12/179 (6.7) | 3/179 (1.7) | 15/358 (4.2) |
| Participants with ≥1 intervention-related safety event, n/N (%) | 10/179 (5.6) | 6/179 (3.4) | 16/358 (4.5) |
| **Event-level outcomes** |  |  |  |
| Total treatment-emergent safety events | 385 | 334 | 719 |
| Events per randomised participant, mean | 2.15 | 1.87 | 2.01 |
| Events per affected participant, mean | 2.79 | 2.32 | 2.55 |
| Adverse events | 362 | 325 | 687 |
| Adverse reactions | 11 | 6 | 17 |
| Serious adverse events | 12 | 3 | 15 |
| Serious adverse reactions | 0 | 0 | 0 |
| Suspected unexpected serious adverse reactions | 0 | 0 | 0 |

*Note: Treatment-emergent safety events include adverse events, adverse reactions, and serious adverse events recorded after randomisation. Pre-treatment emergent events were excluded. Participant-level percentages are calculated using the intention-to-treat population as the denominator. Event-level rows report counts unless otherwise specified.*

*Table S4. Classification and adjudication characteristics of treatment-emergent safety events by randomised group, intention-to-treat population*

| **Characteristic** | **Lifestyle therapy**  **n = 385 events** | **Psychotherapy**  **n = 334 events** | **Total**  **n = 719 events** |
| --- | --- | --- | --- |
| **Event type** |  |  |  |
| Adverse event | 362 (94.0) | 325 (97.3) | 687 (95.5) |
| Adverse reaction | 11 (2.9) | 6 (1.8) | 17 (2.4) |
| Serious adverse event | 12 (3.1) | 3 (0.9) | 15 (2.1) |
| **Severity** |  |  |  |
| Mild | 269 (69.9) | 254 (76.0) | 523 (72.7) |
| Moderate | 83 (21.6) | 68 (20.4) | 151 (21.0) |
| Severe | 32 (8.3) | 11 (3.3) | 43 (6.0) |
| Life-threatening | 1 (0.3) | 1 (0.3) | 2 (0.3) |
| **Relatedness to intervention** |  |  |  |
| Not related | 362 (94.0) | 323 (96.7) | 685 (95.3) |
| Unlikely | 12 (3.1) | 5 (1.5) | 17 (2.4) |
| Possible | 6 (1.6) | 0 (0.0) | 6 (0.8) |
| Probable | 1 (0.3) | 1 (0.3) | 2 (0.3) |
| Highly probable | 4 (1.0) | 5 (1.5) | 9 (1.3) |
| **Relatedness to study procedures** |  |  |  |
| Not related | 377 (97.9) | 328 (98.2) | 705 (98.1) |
| Unlikely | 6 (1.6) | 0 (0.0) | 6 (0.8) |
| Possible | 1 (0.3) | 3 (0.9) | 4 (0.6) |
| Probable | 1 (0.3) | 1 (0.3) | 2 (0.3) |
| Highly probable | 0 (0.0) | 2 (0.6) | 2 (0.3) |
| **Expectedness among assessed events** |  |  |  |
| Expected | 8/12 (66.7) | 8/11 (72.7) | 16/23 (69.6) |
| Unexpected | 4/12 (33.3) | 3/11 (27.3) | 7/23 (30.4) |

*Note. Values are n (%) unless otherwise stated. Percentages are calculated using treatment-emergent event totals within each randomised group, except for expectedness, which is reported as n/N (%) among events requiring expectedness assessment. Expectedness was assessed for events judged to have a plausible causal relationship to the intervention or study procedures.*

*Table S5. Treatment-emergent safety event–System Organ Class pairs by randomised group, intention-to-treat population*

| **MedDRA System Organ Class** | **Lifestyle therapy**  **n = 476 event–SOC pairs** | **Psychotherapy**  **n = 416 event–SOC pairs** | **Total**  **n = 892 event–SOC pairs** |
| --- | --- | --- | --- |
| Psychiatric disorders | 199 (41.8) | 191 (45.9) | 390 (43.7) |
| Infections and infestations | 53 (11.1) | 49 (11.8) | 102 (11.4) |
| General disorders and administration site conditions | 38 (8.0) | 32 (7.7) | 70 (7.8) |
| Gastrointestinal disorders | 30 (6.3) | 32 (7.7) | 62 (7.0) |
| Musculoskeletal and connective tissue disorders | 25 (5.3) | 19 (4.6) | 44 (4.9) |
| Nervous system disorders | 26 (5.5) | 16 (3.8) | 42 (4.7) |
| Social circumstances | 18 (3.8) | 22 (5.3) | 40 (4.5) |
| Injury, poisoning and procedural complications | 24 (5.0) | 6 (1.4) | 30 (3.4) |
| Metabolism and nutrition disorders | 17 (3.6) | 10 (2.4) | 27 (3.0) |
| Respiratory, thoracic and mediastinal disorders | 10 (2.1) | 5 (1.2) | 15 (1.7) |
| Reproductive system and breast disorders | 8 (1.7) | 6 (1.4) | 14 (1.6) |
| Cardiac disorders | 5 (1.1) | 3 (0.7) | 8 (0.9) |
| Investigations | 4 (0.8) | 4 (1.0) | 8 (0.9) |
| Vascular disorders | 2 (0.4) | 5 (1.2) | 7 (0.8) |
| Surgical and medical procedures | 4 (0.8) | 3 (0.7) | 7 (0.8) |
| Endocrine disorders | 4 (0.8) | 3 (0.7) | 7 (0.8) |
| Skin and subcutaneous tissue disorders | 1 (0.2) | 3 (0.7) | 4 (0.4) |
| Eye disorders | 1 (0.2) | 2 (0.5) | 3 (0.3) |
| Neoplasms benign, malignant and unspecified (inc cysts and polyps) | 2 (0.4) | 1 (0.2) | 3 (0.3) |
| Blood and lymphatic system disorders | 1 (0.2) | 1 (0.2) | 2 (0.2) |
| Immune system disorders | 2 (0.4) | 0 (0.0) | 2 (0.2) |
| Renal and urinary disorders | 1 (0.2) | 1 (0.2) | 2 (0.2) |
| Ear and labyrinth disorders | 0 (0.0) | 2 (0.5) | 2 (0.2) |
| Pregnancy, puerperium and perinatal conditions | 1 (0.2) | 0 (0.0) | 1 (0.1) |

*Note. Values are n (%) unless otherwise stated. Counts represent unique safety event–System Organ Class pairs. One treatment-emergent safety event could map to more than one MedDRA System Organ Class; therefore, System Organ Class counts exceed the total number of treatment-emergent safety events. Percentages are calculated using total event–SOC pairs within each randomised group. MedDRA = Medical Dictionary for Regulatory Activities; SOC = System Organ Class.*
