## Supplementary Table 7 for "Lifestyle therapy versus cognitive behavioural therapy for adults with mood disorders: a randomised non-inferiority trial"

*Table S6. Protocol deviations for total sample and intervention arms*

| **Protocol deviation category** | **Examples** | **Total Sample** | **Lifestyle** | **Psychotherapy** |
| --- | --- | --- | --- | --- |
| Attendance deviations | Planned/unplanned non-attendance, late arrival, early departure | 516 | 271 | 245 |
| Assessment deviations | Surveys not completed, missing questionnaires, additional SCID modules conducted (*n*=3) | 464 | 317 | 147 |
| Visit schedule deviations | Assessments or session visits conducted outside protocol window | 85 | 40 | 45 |
| Technology/administrative deviations | Videoconferencing or recording failures, email failure | 29 | 1 | 28 |
| Program delivery deviations | Sessions exceeding planned duration; altered content | 22 | 4 | 18 |
| Unblinding events | Participant unblinding of research staff | 15 | 8 | 7 |
| Privacy/confidentiality deviations | Participants sharing contact details; participant email addresses visible in calendar invitations or group email communications | 10 | 0 | 10 |
| Staffing/training deviations | Facilitator illness, absence, or substitution | 7 | 0 | 7 |
| Other | Randomisation below group minimum numbers | 1 |  |  |
| **Total** |  | 1149 |  |  |
